# Molecular Epidemiology, Diagnostics and Mechanisms of Antibiotic Resistance in *Mycobacterium tuberculosis* complex in Africa: A Systematic Review of Current Reports

**DOI:** 10.1101/19008052

**Authors:** John Osei Sekyere, Melese Abate Reta, Nontuthuko Excellent Maningi, Petrus Bernard Fourie

## Abstract

**Background:** Tuberculosis (TB) remains a main global public health problem. However, a systematic review of TB resistance epidemiology in Africa is wanting.

**Methods:** A comprehensive systematic search of PubMed, Web of Science and ScienceDirect for English research articles reporting on the molecular epidemiology of *Mycobacterium tuberculosis* complex resistance in Africa from January 2007 to December 2018 was undertaken.

**Results and conclusion:** Qualitative and quantitative synthesis were respectively undertaken with 232 and 186 included articles, representing 32 countries. TB monoresistance rate was highest for isoniazid (59%) and rifampicin (27%), particularly in Zimbabwe (100%), Swaziland (100%), and Sudan (67.9%) whilst multidrug resistance (MDR) rate was substantial in Zimbabwe (100%), Sudan (34.6%), Ivory Coast (24.5%) and Ethiopia (23.9%). Resistance-conferring mutations were commonly found in *katG* (n=3694), *rpoB* (n=3591), *rrs* (n=1272), *inhA* (n=1065), *pncA* (n=1063) and *embB* (n=705) in almost all included countries: S**315**G/I/N/R/T, V**473**D/F/G/I, Q**471**H/Q/R/Y, S**303**C/L etc. in *katG;* S**531**A/F/S/G, H**526**A/C/D/G, D**516**A/E/G etc. in *rpoB;* A**1401**G, A**513**C etc. in *rrs;* C**15**T, G**17**A/T, -A**16**G etc. in *inhA;* Ins**456**C, Ins **172**G, L**172**P, C**14**R, Ins**515**G etc in *pncA*. Commonest lineages and families such as T (n=8139), LAM (n=5243), Beijing (n=5471), Cameroon (n=3315), CAS (n=2021), H (n=1773) etc., with the exception of T, were not fairly distributed; Beijing, Cameroon and CAS were prevalent in South Africa (n=4964), Ghana (n=2306), and Ethiopia/Tanzania (n=799/635) respectively. Resistance mutations were not lineage-specific and sputum (96.2%) were mainly used for diagnosing TB resistance using the LPA (38.5%), GeneXpert (17.2%), whole-genome sequencing (12.3%) and PCR/amplicon sequencing (9%/23%). Intercountry spread of strains were limited while intra-country dissemination was common. TB resistance and its diagnosis remain a major threat in Africa, necessitating urgent action to contain this global menace.

## 1. Introduction

As one of the oldest diseases of mankind, it is still surprising and disturbing that Tuberculosis (TB) continues to remain the leading cause of morbidity and mortality from a single aetiological agent worldwide (1–3), with an estimated 10.4 million active TB cases and nearly 1.6 million deaths in 2018 (4). Disturbingly, poorer and overpopulated regions with overburdened healthcare systems such as Africa and South-East Asia comprised 82% of TB deaths among HIV-negative people (5), showing that high TB incidence cases occurred in low-income and resource-limited countries (6–8). Particularly in Africa where the TB incidence rate is around 25% (5), the situation is worsened by social conditions, the HIV epidemic, weak healthcare systems, scarce laboratories, and the emergence of multidrug-resistant-TB (MDR-TB) (7, 9, 10).

Effective TB prevention and control, diagnosis and treatment, particularly of drug-resistant (DR)-TB, needs a large proportion of national budgets [15]. Moreover, DR-TB also affects economic activity as most TB patients are economically active adults with dependents [14]. Dheda *et al*. (2014) revealed that in certain African countries such as South Africa, where DR-TB accounts for less than 3% of all TB cases, over a third of the national TB budget is sapped up by DR-TB alone, which is unmaintainable and threatens to weaken nationwide TB programmes [14, 15]. Similarly, a cost analysis study in South Africa by Pooran et al. (2013) revealed that smear-positive drug sensitive-TB (DS-TB) costed $191.66 per case, whereas a smear-negative and retreatment cases costed $252.54 and $455.50, respectively, making the latter two more expensive [15].

The TB menace in Africa is compounded by the burgeoning prevalence of DR-TB (11), MDR-TB and extensively drug-resistant (XDR-) TB (12), which are impossible to treat with first line drugs and result in higher mortalities, increased medication costs and drug-associated toxicities (13–15). Delays in diagnosis and treatment of TB, previous exposure to anti-TB drugs (2, 11, 16, 17), inappropriate drug regimens (18–20), poor adherence to prescribed regimen (19), and primary infections with DR-TB strains are the main contributing risk factors for the emergence and dissemination of DR-TB (21, 22). The emergence of DR-

TB has necessitated the development of rapid and simple diagnostic methods to detect drug resistance and evolving resistance mechanisms in the *M. tuberculosis* (*M. tb*) genome (2, 12). Evidently, early and accurate detection of DR-TB enhances appropriate treatment and informs tailored interventions to break the TB transmission chain (2). However, the dearth of advanced molecular diagnostic tools, well-equipped laboratory infrastructure and trained personnel in most resource-limited countries in Africa, limits the detection of DR-TB to the use of non-molecular techniques that are time-consuming and have limited efficiency (23, 24).

Thus, DR-TB is perceived after persistent and/or exacerbating TB symptoms several weeks or months after treatment with first-line antibiotics in such settings, consequently selecting and proliferating MDR and XDR-TB (20, 25). Abdelaal *et al*. (2009) revealed that DR-TB was high (70%) in previously treated patients than in newly infected patients (30%) (2) whilst Affolabi *et al*. (2017) confirmed that patients with a history of TB chemotherapy are more likely to develop MDR-TB strains than newly treated patients (16). These findings buttress the need for efficient, simpler and cheaper TB and DR-TB diagnostics for a comprehensive containment of the TB menace.

The number of antibiotics to which *M. tb* is resistant to is also used to classify the level of resistance as mono resistance, MDR, and XDR. *M. tuberculosis* that are resistant to only one of the first-line drugs such as isoniazid (INH), rifampicin (RIF), ethambutol (EMB) and pyrazinamide (PZA) are described as mono-resistant whilst those that are resistant to at least both INH and RIF are defined as MDR. MDR-TB with additional resistance to a fluoroquinolone (FQ) and at least one of the three injectable second-line anti-TB drugs such as amikacin (AMK), kanamycin (KAN) and capreomycin (CAP) is classified as XDR-TB (5, 11, 20, 26, 27).

Drug resistance in *M. tb* is mediated through various mechanisms. Firstly, the hydrophobic mycolic acid-saturated architecture of the *M. tb* cell wall reduces permeability to several antibiotics (20, 27–29). Secondly, mutations in the hotspot regions of genes encoding drug target proteins or enzymes result in drug resistance (27, 30). Finally, active efflux of antibiotics and expression of drug-deactivating enzymes protect the bacilli against antibiotics (27, 28). In this review, the epidemiology of resistance-conferring mutations i.e., deletions, insertions, frameshift or point mutations, described in genes mediating resistance to first-line antitubercular drugs viz., INH, RIF, PZA, EMB, and streptomycin (STM) (18, 31–37), as well as for most second-line antitubercular drugs such as ethionamide (ETM), FQ, CAP, KAN, and AMK (36–38) are described. Unlike in Gram-Positive and Gram-Negative bacterial species, horizontal acquisition of resistance genes are unknown in TB (20, 39). However, Mehra et al. (2016), recently reported on the presence of drug resistance ABC (efflux) transporters and important virulence factors/genes such as the ESX-1 transport machinery, including ESAT-6 and CFP-10, as well as ESX-5 regions of the type-VII secretion system (T7SS) on a genomic island in *M. tuberculosis* genomes, suggesting horizontal gene transmission.

Molecular epidemiology analysis decodes strain transmission dynamics, risk factors for recent transmission, the occurrence of mixed *M. tb* infections (40), disease severity and drug resistance (41), as well as TB strains related to outbreaks (42). There are currently seven *M. tb* lineages, comprised of several sub-lineages and families, with region-specific prevalence rates and varying virulence capacities (43). Specifically, the Beijing family has been implicated in several studies as having a better capability for developing MDR (40, 44, 45). Genotyping tools such as spoligotyping, MIRU-VNTR typing, IS*6110* RFLP, and whole-genome sequencing (WGS) are normally used to determine the lineage and transmission routes of *M. tb* in Africa, albeit WGS provides a better typing and evolutionary epidemiology resolution.

### Evidence before this review

Many studies have reviewed the resistance rate of antitubercular drugs, mechanisms of resistance, and distributions of *M. tuberculosis* strains/lineages in distinct African countries, but not throughout Africa. A thorough search of the literature in Pubmed within the last five years for reviews addressing the molecular epidemiology, resistance rates and diagnostics throughout Africa yielded no results, suggesting that this could be the first to review these on the continent.

### Purpose of this review

This review seeks to describe the molecular epidemiology of *M. tb* strains/genotypes per country, their mechanisms of resistance, and mono-resistance, MDR and XDR rates in Africa. Furthermore, we describe herein the molecular diagnostic methods used to identify and characterize antitubercular drug resistance as well as the genotyping methods used to identify *M. tb* lineages/sub-lineages in Africa. It is our aim that these findings shall inform evidence-based public health TB intervention policies as well as the prescription, stewardship and design/development of novel antitubercular medications.

## 2. Methods

### Databases and Search Strategy

A comprehensive literature search was carried out on PubMed, Web of Science, and ScienceDirect electronic databases. English research articles published between January 2007 and December 2018 were retrieved and screened using the following search terms and/or phrases: “molecular epidemiology”; “*Mycobacterium tuberculosis*”; “tuberculosis”; “mechanisms of drug resistance”; “genetic diversity”; “transmission dynamics”; “genotyping”; drug resistance”; “MDR-TB”; “XDR-TB”; “gene mutations”; “first- and second-line anti-TB drug”; “frequency of gene mutation”; and “Africa”. These search words/phrases were further paired with each other or combined with names of each African country, and search strings were implemented using “AND” and “OR” Boolean operators. The search focused on the molecular epidemiology of *M. tb* and/or mechanisms of antitubercular drug resistance.

### Inclusion and Exclusion Criteria

Articles not reporting on *M. tb* genotypes and anti-TB drug resistance mechanisms were excluded. Studies addressing the following were included: [a] molecular epidemiology/genotype of *M. tb* strain*s*; [b] resistance-conferring mutations associated with first and second-line antitubercular drugs; [c] molecular diagnostics; [d] TB research conducted in Africa; [e] studies conducted between January 2007 and December 2018; and [f] studies published in English.

This review was conducted using the Preferred Reporting Items for Systematic Reviews and Meta-Analyses (PRISMA) guidelines (Figure 1). The following data were extracted from the included articles: country; year of study; specimen source(s); genotyping and diagnostic method(s)/techniques used; *M. tb* strains/lineages/sub-lineages; sample size; resistant isolates and resistance rates; frequency of resistance-conferring mutations per gene; antitubercular drug resistance mechanisms (Tables S1-S6).

**Figure 1.**
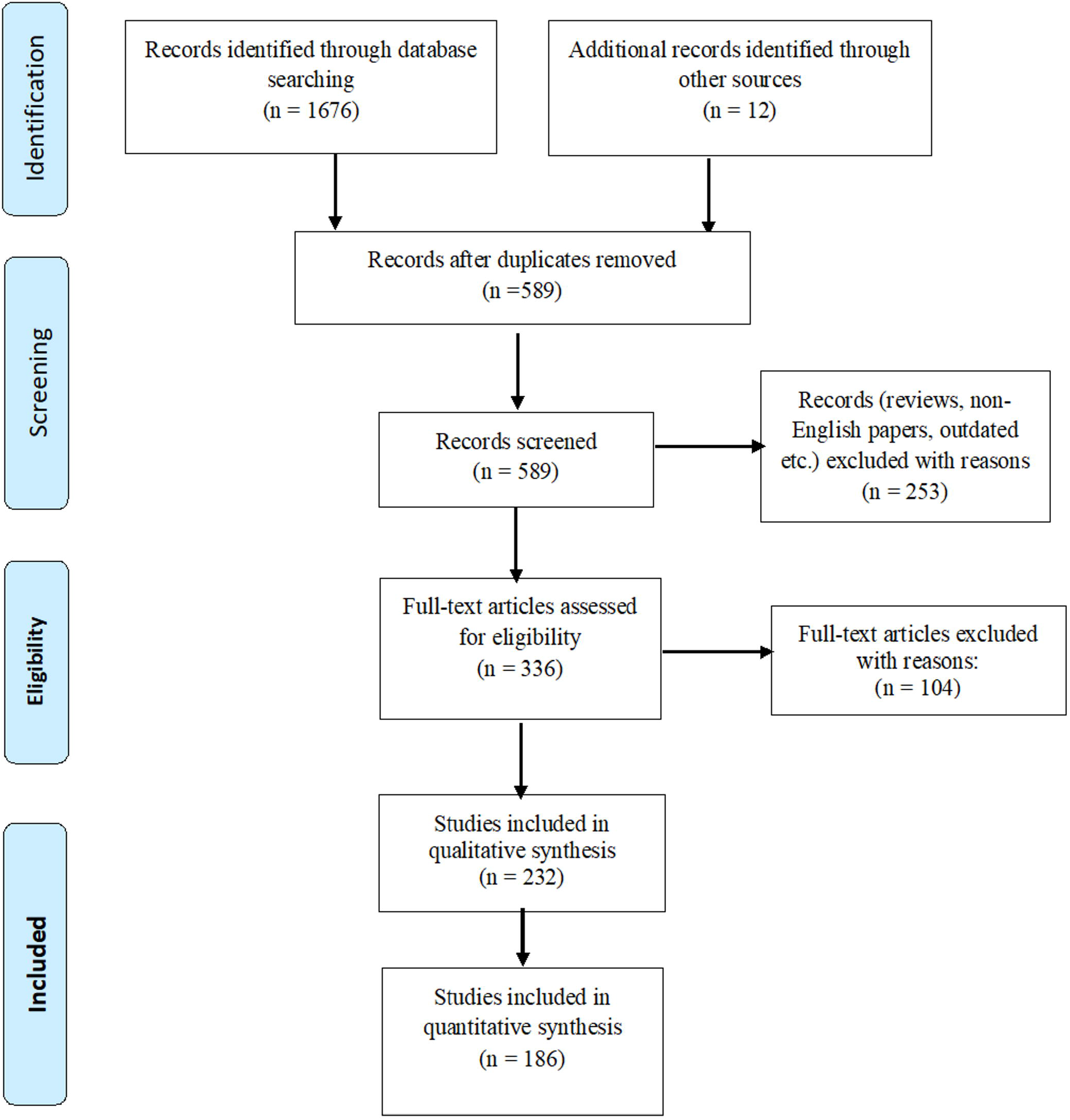
PRISMA –adapted flow diagram showed the results of the search and reasons for exclusion. Adapted from: Moher D, Liberati A, Tetzlaff J, Altman DG, The PRISMA Group (2009). *P*referred *R*eporting *I*tems for *S*ystematic Reviews and *M*eta-*A*nalyses: The PRISMA Statement. *PLoS Med* 6(7): e1000097. doi:10.1371/journal.pmed1000097.

The rate of resistance to anti-TB drugs was evaluated per country to identify regions with the lowest and highest burden of mono-resistance, MDR and XDR rates. Resistance mutations distribution per gene were examined to identify countries with the most resistance as well as the mutation rate per locus of each gene. The distribution of *M. tb* lineages/sub-lineages and families were determined per country. The association of *M. tb* lineages with specific gene mutations were assessed.

### Phylogenomics

Genomes of *M. tb* isolated from Africa from 1997 to 2019 (n=800 isolates) were downloaded from PATRIC (https://www.patricbrc.org/) and categorized into resistant isolates and mixed (both resistant and non-resistant) isolates (Supplementary dataset). RAxML v8.2 was used to draw the core-genome phylogenetic trees of these strains, using *Streptococcus mitis* M3-4 as an outgroup/reference strain (46). The trees were subsequently annotated with Figtree (http://tree.bio.ed.ac.uk/software/figtree/). Isolates of the same country were given the same label colours. Clades and subclades were highlighted with the same colours to show their closer relationship.

### Statistical analysis

Microsoft Excel® 365 were used to analyze the resistance rate and frequency data in this study using the raw data extracted from the included articles. The resistance rates per antibiotic per country were calculated by dividing the total resistant isolates by the total isolates for which antibiotic sensitivity was determined.

## 3. Results

### Characteristics of included studies

As described in Figure 1, 1688 potential research articles were documented: 1598 articles from PubMed, 41 articles from Web of Science, 37 articles from ScienceDirect, and 12 articles from other sources obtained through manual search. Of the total, 589 were non-duplicated and subjected to further evaluation, 253 were excluded based on the title and abstract evaluation whilst 336 were retained on detailed full-text review. After full-text evaluation, 232 studies on the molecular epidemiology and/or mechanisms of antitubercular drug resistance were used for qualitative review analysis, and 186 studies were used for quantitative analysis (Tables S1-S6).

The 186 studies were from 32 out of 54 (59.3%) countries in Africa, and these included Algeria (n=1 study, 126 isolates), Angola (n=1 study, 89 isolates), Benin (n=2 studies, 294 isolates), Burkina Faso (n=2 studies, 183 isolates), Botswana (n=1 study, 260 isolates), Cameroon (n=4 studies, 1334 isolates), Chad (n=1 study, 311 isolates), Congo (n=2 studies, 120 isolates), Ivory Coast (n=3 studies, 261 isolates), Djibouti (n=3 study, 529 isolates), Egypt (n=5 studies, 697 isolates), Ethiopia (n=36 studies, 7 338 isolates), Gabon (n=1 study, 124 isolates), Ghana (n=7 studies, 7 527 isolates), Guinea (n=1 study, 184 isolates), Guinea-Bissau (n=2 studies, 514 isolates), Madagascar (n=1 study, 158 isolates), Malawi (n=3 studies, 2054 isolates), Mali (n=2 studies, 571 isolates), Morocco (n=7 studies, 1966 isolates), Mozambique (n=6 studies, 884 isolates), Nigeria (n=8 studies, 1984 isolates), Rwanda (n=2 studies, 221 isolates), Sierra Leone (n=2 studies, 194 isolates), South Africa (n=46 studies, 18 635 isolates), Swaziland (n=2 studies, 203 isolates), Sudan (n=5 studies, 469 isolates), Tanzania (n=9 studies, 1 756 isolates), Tunisia (n=4 studies, 1 701 isolates), Uganda (n=13 studies, 3 873 isolates), Zambia (n=2 studies, 395 isolates) and Zimbabwe (n=2 studies, 338 isolates) (Table S1).

There were ∼55, 924 *M. tb* isolates from the 232 studies that were isolated from different specimen sources, including sputum specimens (85.5%, n=51419 isolates), fine needle aspirates (FNA) (4.8%, n=1095 isolates), fine needle aspirates and sputum (3.8%, n=2779 isolates), and gastric lavage and sputum (2.2%, n=232 isolates) (Table 1). The molecular epidemiology of *M. tb* strains/lineages in Africa were found in 126 studies whilst 95 studies reported on only mechanisms of resistance (gene mutations) associated with first and second-line anti-TB drugs (Table S2-S4). As well, 28 studies reported on *M. tb* strains/lineages associated with specific mutations conferring resistance to first and second-line anti-TB drugs in Africa (Table S6). Fifty-three studies reported on the target gene or whole-genome sequencing analysis after performing phenotypic drug susceptibility testing with the conventional proportion (agar dilution) methods on Lowenstein-Jensen agar medium and BACTEC MGIT 960 and 460 systems (Tables 2 & S5). The least sample size per study were observed in Ivory Coast (n=2), South Africa (n=2) and Uganda (n=2); the largest sample sizes were observed in South Africa (n=4667). A molecular characterization of *M. tb* strains and/or mechanisms of drug resistance (gene mutations) associated with drug resistance were performed for all included studies (Table S1-S6).

**Table 1.** Specimen sources used for molecular epidemiology and mechanisms of drug resistance diagnosis across African countries: January 2007-December 2018.

| Specimen source(s) | Number of studies(n) | Percent (%) | Total isolates | Percent (%) |
| --- | --- | --- | --- | --- |
| Animal bronchial washes and human sputum | 1 | 0.54 | 2 | 0.004 |
| Biopsy specimens | 1 | 0.54 | 56 | 0.10 |
| Biopsy, lymph node aspirates and blood specimens | 1 | 0.54 | 95 | 0.17 |
| Cattle milk | 1 | 0.54 | 11 | 0.02 |
| Fine needle aspirates (FNA) | 9 | 4.8 | 1095 | 1.96 |
| Pleural, peritoneal and synovial fluids | 1 | 0.54 | 59 | 0.11 |
| Sputum | 159 | 85.5 | 51419 | 91.94 |
| Gastric lavage and sputum | 4 | 2.2 | 232 | 0.42 |
| Fine needle aspirates and sputum | 7 | 3.8 | 2779 | 4.97 |
| Sputum (induced), bronchial wash, bronchial aspirations, pleural fluid and gastric liquid | 1 | 0.54 | 168 | 0.3 |
| Tuberculous lesions from Cattle and buffaloes | 1 | 0.54 | 8 | 0.014 |
| **Total** | **186** | **100%** | **55,924** | **100%** |

**Table 2.** *M. tuberculosis* molecular diagnostic methods used in Africa January 2007 - December 2018

| Phenotypic/Molecular diagnostic assays | Number of studies (n=186) | Percentage of studies (%) |
| --- | --- | --- |
| **Phenotypic assays** |  |  |
| Agar-dilution/modified proportion method on LJ | 27 | 14.5 |
| BACTEC MGIT 960 system | 18 | 9.7 |
| BacT/ALERT 3D/460 systems | 6 | 3.2 |
| MIC test/Sensititre MYCOTB plate (TREK Diagnostics, Cleveland, OH, USA) | 2 | 1.1 |
| **Molecular assays** |  |  |
| Hain's (GenoType®MTBDR) line probe assay (LPA) | 47 | 25.3 |
| PCR and amplicon sequencing (Applied Biosystems ABI sequencers) | 39 | 17.1 |
| GeneXpertMTB/RIF assay | 21 | 11.3 |
| Whole genome sequencing | 15 | 8.1 |

### Rate of drug resistance and isolates source(s)

From the ∼55 924 total isolates, drug sensitivity tests (DST) were done for 24 836 isolates, of which 23 959 isolates were resistant to at least one antibiotic, resulting in an overall resistance rate of 96.5% (Table S5). The mono-resistance (to at least one antibiotic) and MDR rate per country is shown in Figure 2 and Tables S3 and S5. Mono-resistance and MDR to any of the anti-TB drugs was found in only 25 countries out of 32 countries included in this review, with INH and RIF having the highest monoresistance rates (Fig. 2A-2C). MDR rate was highest in Zimbabwe (100%), Swaziland (100%) and Guinea Bissau (88.9), albeit South Africa had the highest number (n=4579) of MDR isolates. MDR rates were relatively low in other countries (Fig. 2D). The rate of patients with MDR and XDR-TB strains in Africa were 14.5% (3 598 MDR isolates), and 3.1% (781 XDR isolates), respectively. However, all the XDR isolates were from South Africa.

**Figure 2.**
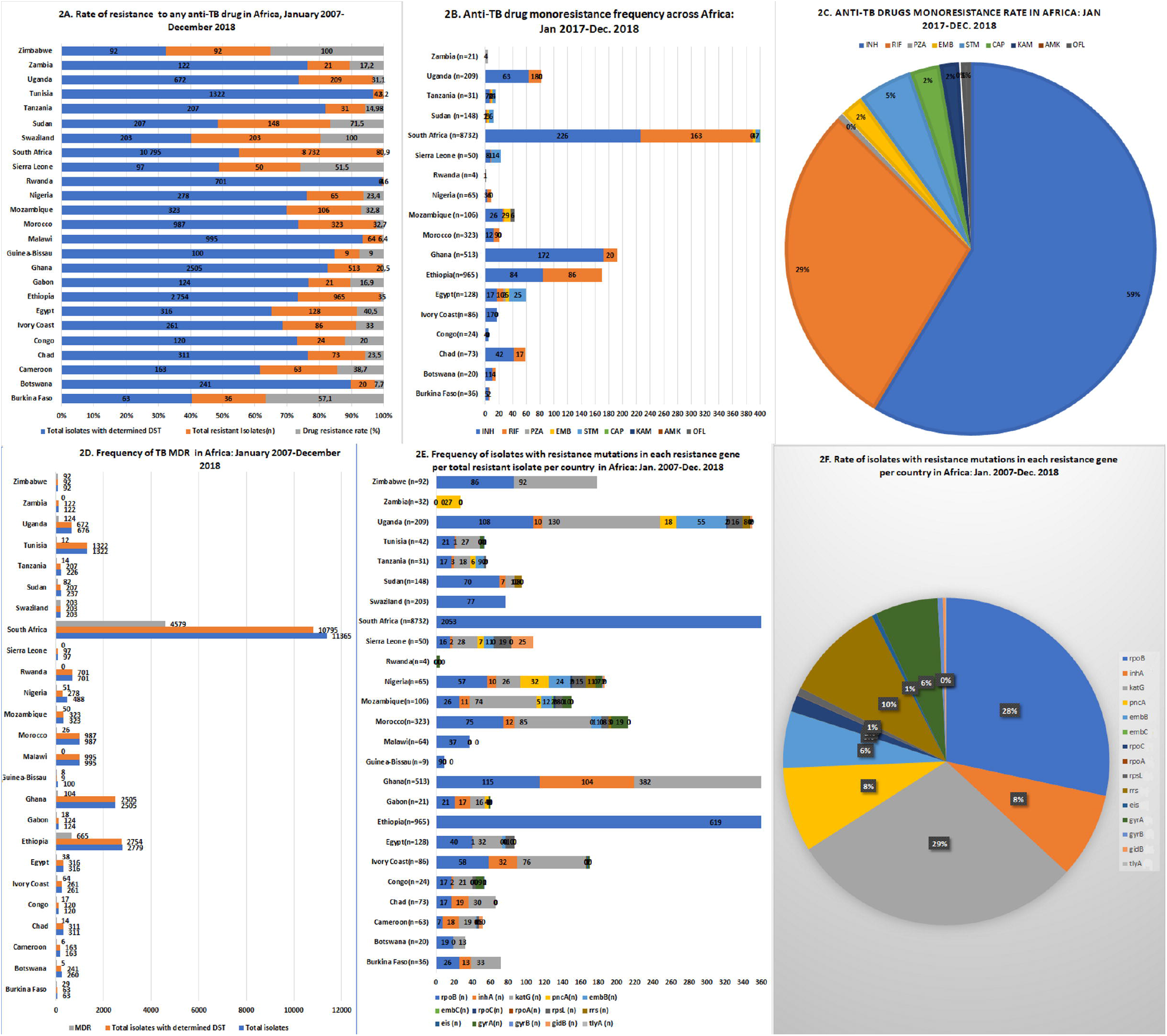
Drug resistance rate and mechanisms of resistance (resistance determinants) of *M. tuberculosis* in Africa, January 2007 to December 2018. (**A**) Rate of resistance to any anti-TB drug in Africa. Total resistance rate (%) of *M. tuberculosis* isolates across African countries, calculated per total isolates in each country; (n=number of resistant isolates). (**B**) Anti-TB drug monoresistance per country in Africa. Each antibiotic’s mono-resistance rate is calculated per total isolates in each country with known resistance profile (n=total number of isolates). (**C**) Total anti-TB drug monoresistance rate per antibiotic in Africa. The resistance rate of each antibiotic is calculated per total *M. tuberculosis* isolates with known resistance in Africa (n=number of resistant isolates per antibiotic). **(D**) Multidrug resistance rate (%) of *M. tuberculosis* across African countries; the number of MDR isolates is calculated per total isolates with known resistance in each country (n=number of MDR isolates). (**E**) Frequency of isolates with resistance mutations in each resistance gene per country in Africa.; the frequency of mutations per each gene is calculated from total isolates in each country (n=number of resistant isolates). (**F**) Rate (%) of genes having resistance-conferring mutations in African *M. tuberculosis* isolates; it is calculated by dividing isolates with resistance genes by all resistant isolates in Africa (n=number of isolates with particular gene mutation).

In almost all the 25 countries, resistance mutations were found in mainly the *katG* (29%), *rpoB* (28%), *rrs* (10%) and *inhA* (8%) genes, with *pncA* (8%) mutations being commonly found in Zambia, Uganda, Tanzania, Sierra Leone, Nigeria and Gabon. *embB* (6%) was common in Uganda, Tanzania, Sierra Leone, Nigeria, Mozambique, Morocco and Egypt. Resistance mutations in *gyrA* (6%) were also common in few countries such as Tunisia, Rwanda, Nigeria, Mozambique, Morocco, and Congo (Fig. 2E and 2F).

Within each of these resistance genes, there were common mutations that were found in isolates from all countries (Figures 3–5; Tables S2-S4; FIG. 4). For instance, in *rpoB*, S**531**A/F/L/S/Q/T/W/Y/STOP, H**526**A/C/D/G/L/N/P/Q/R/T/Y/del, and D**516**A/E/G/V/Y/del were common, S**315**G/I/N/R/T/L/S/+ahpC-48 and V**473**D/F/G/I/K/M/N/R/S/V/ W/Y/STOP were the commonest in *katG*,, C**15**T, G**17**A/T and T**8**A/C were prevalent in *inhA* and its promoter (*inhA*Pro) whilst M**306**I/LV/Xaa was more pronounced in *embB*. Mutations in the *pncA* gene were varied and spread-out on the gene, with Ins**456**C, L**172**P/G/Ins, K**96**R/STOP and C**14**R being dominant. K**43**R and K**88**R/T on *rpsL*, D**94**A/G/H/N/Y, A**90**L/V, D**94**G and S**95**T on *gyrA* as well as A**1401**G, A**513**C, S**2169**A etc. on *rrs* were the most recurring mutations (Fig. 3). These mutations, particularly in *rpoB, katG* and *inhA*, were found in isolates from almost all the countries, with *pncA* mutations being reported mainly from Gabon, Nigeria, Sierra Leone, South Africa, Tanzania, Uganda and Zambia (Fig. 4–5). Reported mutations on the *rpoA/C, embA/C, gyrB, gidB, eis, ndh, ahpC, tlyA* were relatively few and reported in few countries throughout Africa. For instance, *gibB* was reported in Cameroon, Nigeria, Sierra Leone, South Africa, and Uganda, *eis* was reported in Ivory Coast and South Africa, *ndh* was found in Ghana, *ahpC* was found in Ghana and Uganda, and *tlyA* was found in South Africa (Figure 3–5; Tables S2-S4; FIG. 4).

**Figure 3:**
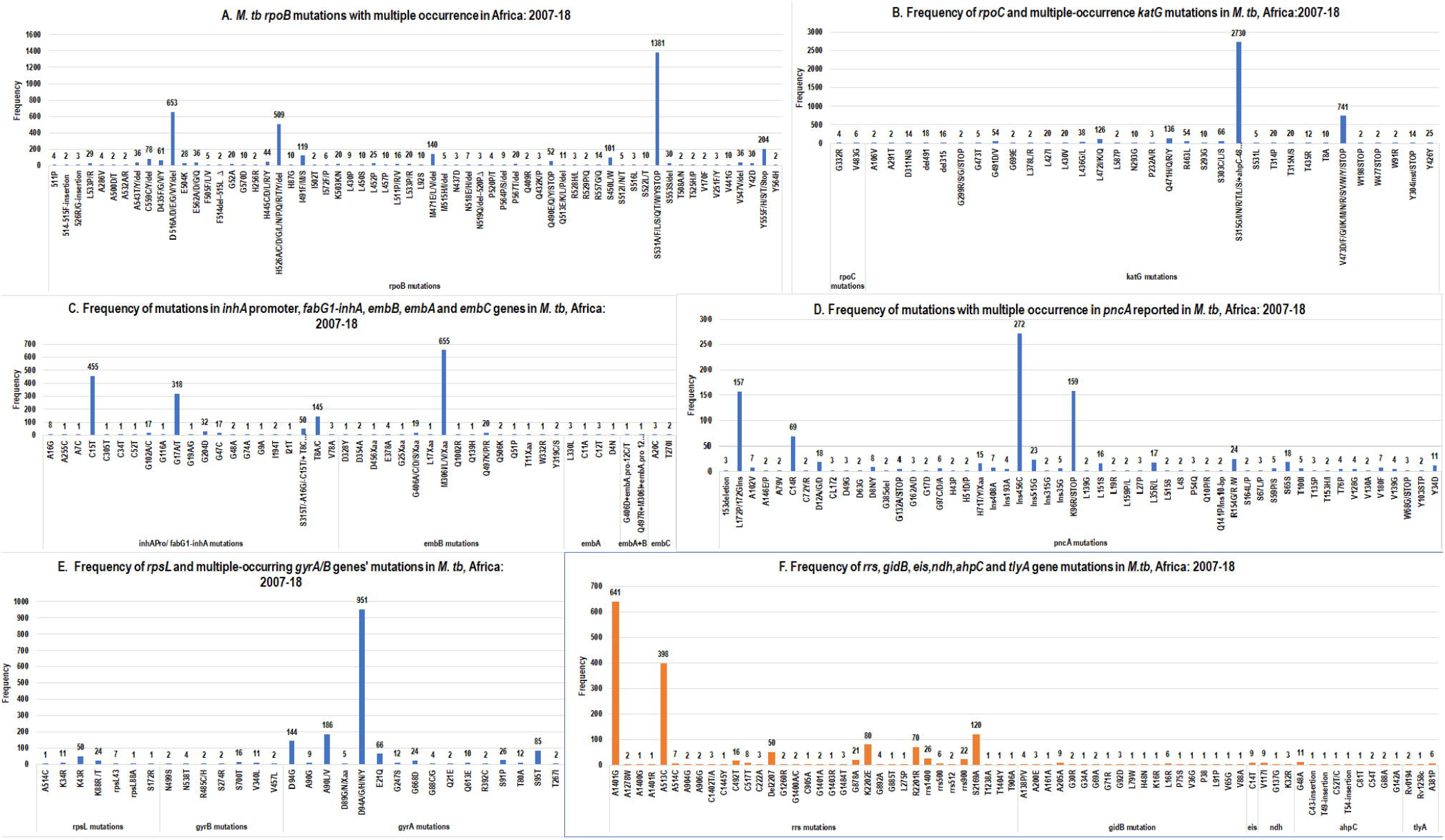
*M. tuberculosis* anti-TB drug resistance mutations across Africa, January 2007-December 2018. The frequencies of each mutation per gene in all isolates across Africa is shown in the image. (**A**) *M. tuberculosis rpoB* mutations with multiple occurrence in Africa. (**B**) Frequency of *rpoC* and multiple occurrence *katG* mutation in *M. tuberculosis* in Africa. (**C**) Frequency of mutations in *inh*A promoter, *fab*G1-*inh*A, *emb*B, *emb*A and *emb*C genes in *M. tuberculosis* in Africa. (**D**) Frequency of mutations with multiple occurrence in *pnc*A reported in *M. tuberculosis* in Africa. (**E**) Frequency of *rps*L and multiple-occurring *gyr*A/B mutations in *M. tuberculosis* in Africa. (**F**) Frequency of *rr*S, *gid*B, *eis, ndh, ahp*C and *tly*A gene mutations in *M. tuberculosis* in Africa.

**Figure 4 (A and B):**
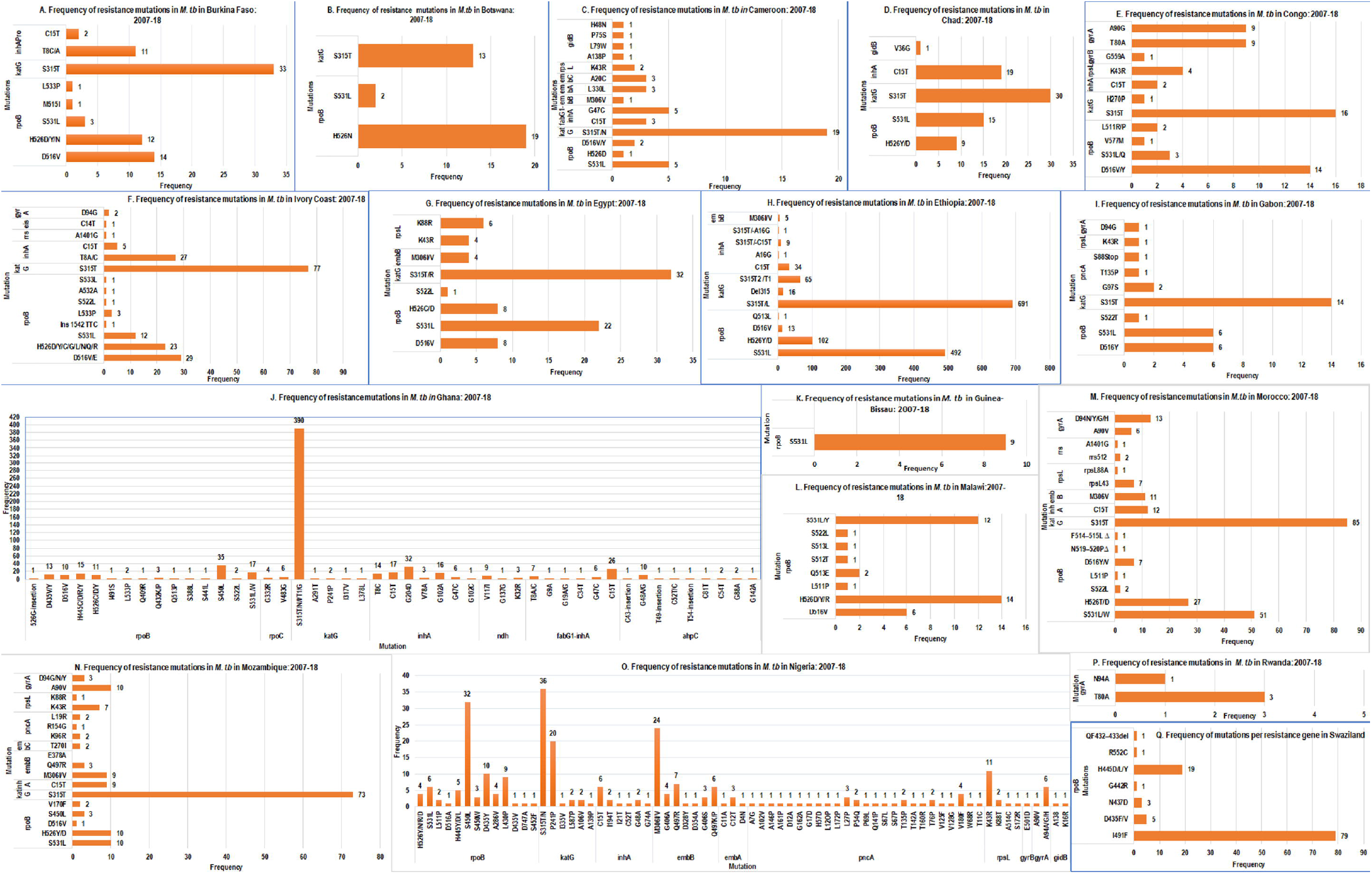
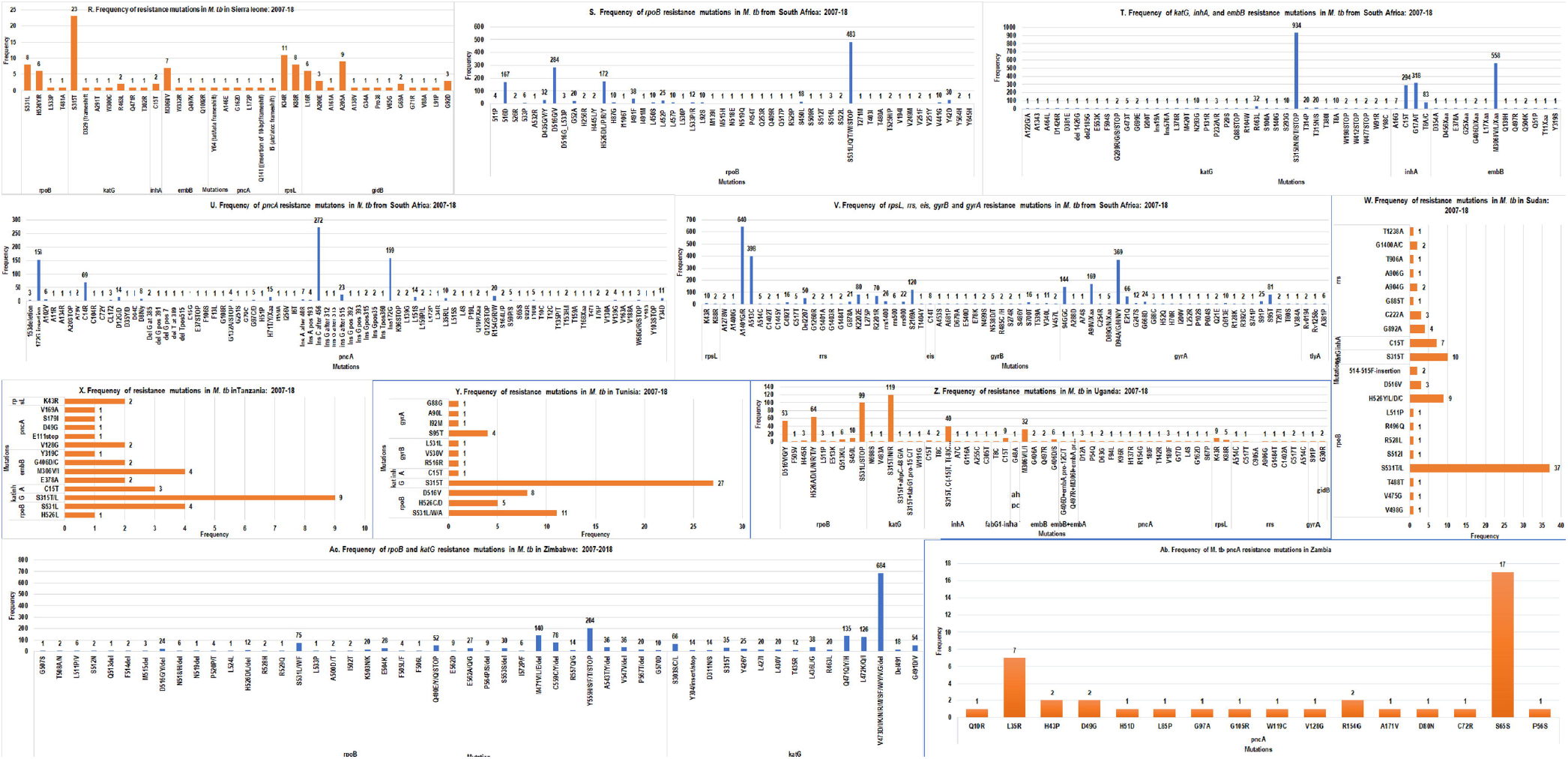
Frequency of resistance mutations in each resistance gene per total resistant isolates per country in Africa, January 2007-December 2018. Frequency of resistance mutations in *M. tuberculosis* in Burkina Faso (**A**), Botswana (**B**), Cameroon (**C**), Chad (**D**), Congo (**E**), Ivory Coast (**F**), Egypt (**G**), Ethiopia (**H**), Gabon (I), Ghana (**J**), Guinea-Bissau (**K**), Morocco (**M**), Mozambique (**N**), Nigeria (**O**), Rwanda (**P**), and Swaziland (**Q**) are shown in 4A. Frequency of resistance mutations in respective genes in *M. tuberculosis* in Sierra Leone (**R**), South Africa (**S, T, U and V**), Sudan (**W**) Tanzania (**X**), Tunisia (**Y**), Uganda (**Z**), Zimbabwe (**Aa**) and Zambia (**Ab**) are also shown in B.

**Figure 5:**
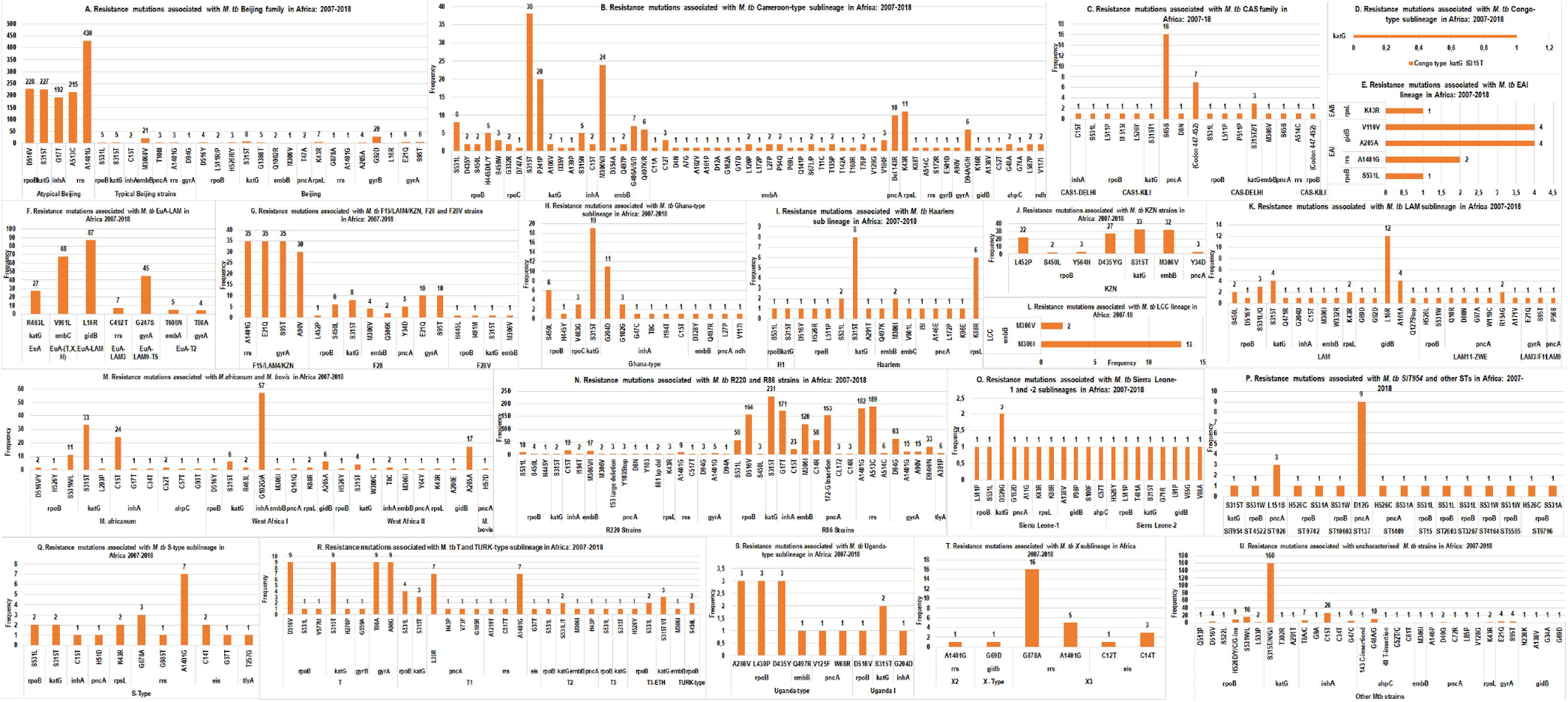
Frequency of resistance mutations associated with *M. tuberculosis* lineages/sublineage or families in Africa, January 2007-December 2018. Resistance mutation associated with *M. tb* Beijing family (**A**), Cameroon-type sublineage (**B**), CAS family (**C**), Congo-type sublineage (**D**), EuA-LAM (**F**), F15/LAM4/KZN, F28 and F28V strains (**G**), Ghana-type sublineage (**H**), Haarlem sublineage (**I**), KZN strains (**J**), LAM sublineage (**K**), *M*.*bovis* (**M**), R220 and R86 strains (**N**), Sierra Leone 1 and 2 sublineages (**O**), SIT954 and other STs (**P**), S-type sublineages (**Q**), T and TURK-type sublineages (**R**), Uganda-type sublineages (**S**), X-sublineages (**T**), and uncharacterized *M*.*tb* strains (**U**) are respectively shown per chart in the image.

Commonest mutations in each resistance gene per lineage, sub-lineage and families are shown in Figure 6 and Table S6. Lineage or family-specific resistance mutations per gene were difficult to find, particularly for mutations that occurred in smaller frequencies. Particularly, *rpoB* (e.g. S**531**L, D**516**Y, H**526**Y) and *katG* (S**315**T) mutations were found in almost all lineages and families in almost all countries, albeit in very low prevalence in Beijing strains, which were predominantly reported from South Africa; *katG* S**315**T mutation was the only one found in the Congo sub-lineage. Mutations in *rrs*, particularly A**1401**G were very pre-dominant in Beijing, F15/LAM4/KZN and X lineages. G**204**D and G**102**A mutations in *inhA* were commonly associated with the Ghana-type sublineage, which was reported mainly from Ghana whilst the C15T mutation was spread throughout almost all countries and lineages (Figure 5; Table S6). Notably, only T**47**A mutation (n=2) in *pncA* was reported in Beijing strains whilst low frequencies of other diverse *pncA* mutations were found in several other families, particularly in South Africa; the sole mutation reported in *M. bovis* was a single *pncA* mutation (H**57**D). Likewise, *embB* mutations were almost absent in Beijing and CAS strains, but was very common in R86, Cameroon, and KZN strains whilst very low frequencies were found in almost all other families (Fig. 5).

**Figure 6 (A and B):**
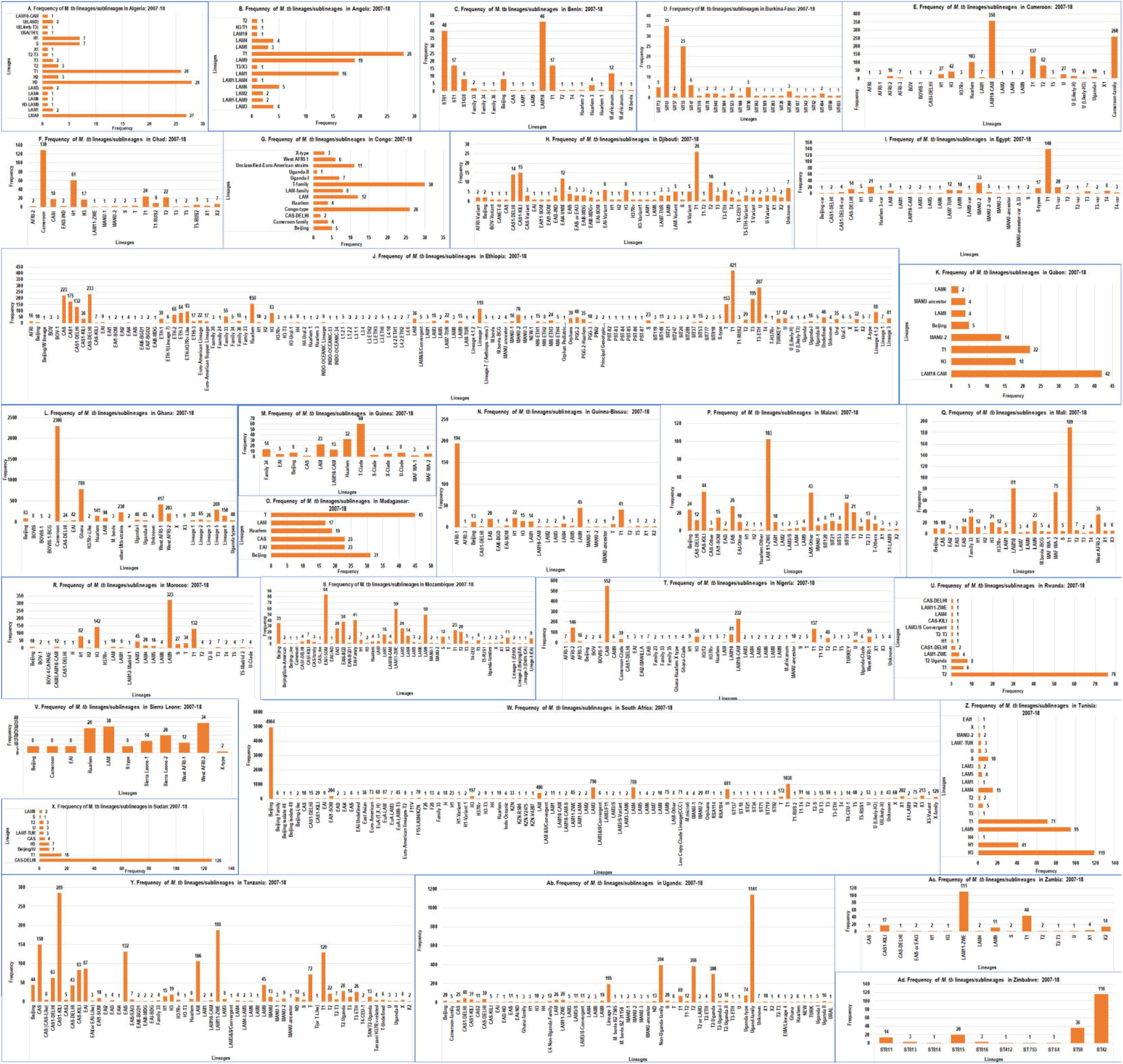
Frequency of *M. tuberculosis* lineages/sublineages or families in Africa, January 2007-December 2018. Frequency of *M. tuberculosis* lineages/sublineage in Algeria (**A**), Angola (**B**), Benin (**C**), Burkina Faso (**D**), Cameroon (**E**), Chad (**F**), Congo (**G**), Djibouti (**H**), Egypt (**I**), Ethiopia (**J**), Gabon (**K**), Ghana (**L**), Guinea (**M**), Guinea Bissau (**N**), Malawi (**P**), Mali (**Q**), Morocco (**R**), Mozambique (**S**), Nigeria (**T**), Rwanda (**U**), Sierra Leone (**V**), South Africa (**W**), Sudan (**X**), Tanzania (**Y**), Tunisia (**Z**), Uganda (**Ab**), Zambia (**Ac**), and Zimbabwe (**Ad**) are shown per chart in the image. The prevalence of each lineage/sublineage differed per country in 6A. 6B shows the distribution of the main lineages per country in Africa. Distinct colour codes depict the various lineages (1–8) with lineage 4 being most common in almost all countries, Lineage 5 and 6 being found in mainly West Africa, and Lineage 7 being reported in only Ethiopia.

Mutations in genes conferring resistance to 2^nd^-line drugs were not as frequent as those in 1^st^-line drugs. Specifically, *gyrA* mutations were very low in Beijing, EuA-T2, and Cameroon strains, high in EuA-LAM9-T5, F28, F15/LAM4/KZN etc. and absent in *M. africanum, M. bovis*, Ghana-type, Haarlem, CAS, Congo, KZN, LCC, SIT954, X, Sierra Leone, Uganda, TURK and S-type strains. Furthermore, *gibB* mutations were prevalent in LAM, including EuA-LAM, and *M. africanum* II strains, but were very low in Cameroon-type, East African Indian (EAI), X-type, Sierra Leone, and uncharacterized strains. Resistance mutations occurred in *gibB* alongside *rpsL* and/or *rrs* in Cameroon, EAI, LAM (EuA-LAM), X, *M. africanum*, Sierra Leone, and uncharacterized strains (Figure 5; Table S6).

### North Africa: Egypt, Morocco, Sudan, Tunisia

#### Egypt

DST was determined for 316 clinical isolates by three studies in Egypt, resulting in a resistance rate of 40.5% (n=128) and a RIF-monoresistance rate of 3.2% (n=10) (Tables S1 & S3), the highest of which was reported by Abdelaal *et al*.(2009) to be 3.9% (n=6) (2). In the same year, Abbadi *et al*.(2009) reported a 1.9% (n=3) RIF mono-resistance rate (47). RIF resistance was mostly mediated by S531L (n=21), D516V (n=8), and H526N/Y (n=8) mutations in the *rpoB* gene (48, 49). Similarly, the mean INH-monoresistance rate in Egypt was found to be 5.4% (n=17), mediated by S315T (n=31) mutation in *kat*G (Table S2).

Abbadi *et al* (2009) reported a PZA-monoresistance rate of 0.6% (n=2), albeit the resistance mechanisms was not characterised (47). However, Bwalya *et al* (2018) in Zambia reported a 3.3% PZA monoresistance rate, with different *pnc*A mutations (50). The MDR rate in Egypt was 12% (n=38) (Table S1 & S5).

EMB-monoresistance rate was 1.9% (n=6), mediated mainly by M306V/I mutations (n=2) (2, 47) in *emb*B (3.1%, n=4) (Tables S1 & S3). Abbadi *et al* (2009) detected the highest STM-monoresistance rate in Egypt, which was 13.5% (n=21), and was mainly mediated by *rps*L gene mutations (55.6%) (47).

#### Morocco

Four studies conducted in Morocco undertook DST of 987 *M. tb* isolates, yielding a resistance rate of 32.7% (n=323), a RIF-monoresistance rate of 0.9% (n=9) and an INH monoresistance rate of 1.2% (n=12) (Tables S1 & S3). S531L (n=48), followed by H526D (n=23) mutations in *rpoB* (23.2%, n=75) predominantly mediated RIF resistance (Tables S1 & S4; FIG. 4). Karimi *et al* (2018) reported an INH monoresistance rate of 17.1% (n=12) (23). The proportion of *kat*G gene mutation was 26.3% (n=85) whilst *inh*A mutation was 3.7% (n=12) (Tables S2 & S3), being respectively mediated by S315T (n=85) and C15T(n=10) (at *inh*A’s promotor region) (Table S4; FIG. 4). The MDR rate in morocco was 2.6% (n=26), and was comparable with the 2016 estimated incidence rate of MDR/RIF resistance cases in the country, which was 1.8/100,000 (5). However, the 2016 national survey reported that the MDR/RIF resistant TB cases among previously treated cases was 11% (5).

#### Sudan

The rate of antibiotics resistance in Sudan, based on three studies, was 67.9% (n=161) (11, 51–53). S531L (n=36) and H526Y(n=5) were the commonest *rpo*B (43.5%, n=70) mutations associated with RIF resistance (Tables S2 & S3). Abdul-Aziz *et al*. (2013) reported an INH monoresistance of 0.4% (n=1) in Sudan (11). Mutations in *kat*G and the *inh*A promotor region was mainly S315T (n=10) and C15T(n=7) respectively, conferring high-level INH resistance. Moreover, the MDR rate in Sudan was 34.6% (n=82) (Tables S1). The highest MDR rate, 100% (n=49), was detected in the year 2013 (53).

#### Tunisia

Three studies in Tunisia, encompassing 1322 clinical isolates, resulted in a 3.2% (n=42) resistance rate. RIF monoresistance rate was not reported in any of the studies. D516V (n=8) and S531W (n=4) mutations in *rpoB* (50%, n=21) commonly mediated RIF resistance (Tables S1-S3). This Tunisian result was lower than the finding detected in Egypt 40.5% (2, 47, 54), Morocco 32.7% (23, 55–57), Sudan 67.9% (11, 51, 53), Mozambique 32.8% (58, 59); but higher than the report from Rwanda 0.6% (60) (Figure 2a). Comparatively, the MDR rate in Tunisia was very low (Tables S1-S3).

### West Africa: Burkina Faso, Ghana, Guinea Bissau, Ivory Coast, Nigeria, Sierra Lone

#### Burkina Faso

A single study by Miotto *et al* (2009), using the MTBDR*plus* on 63 isolates, reported a resistance rate of 57.1% (n=36), and a RIF and INH monoresistance rate of 3.2% (n=2) and 7.9% respectively (Tables S1-S3). D516V (43.8%), followed by H526D (15.6%), H526Y (15.6%), and S531L (9.4%) *rpo*B mutations conferred RIF resistance in 72.2% (n=26) of isolates while mutations in *kat*G, found in 86.8% (n=33) of isolates, and mutation in *inh*A, found in 34.2% (n=13) of strains conferred INH resistance (Tables S4; FIG. 4). The MDR rate was 46.0% (n=29), which is higher than the 2016 estimated incidence of 2% MDR/RIF resistant-TB in the country (1.6/100,000) and the 2017 national survey report of 14% MDR/RIF resistant-TB case among new and previously treated cases (5).

#### Ghana

The overall resistance rate in Ghana, based on 2505 *M. tb* isolates in four studies, was 20.5% (n=513), which is higher than those of Tunisia (3.2%), Malawi (6.4%), Rwanda (0.6%), Guinea Bissau (9%), Nigeria (13.3%) and Gabon (16.9%), but lower than those reported form other African countries (Tables S1-S3). RIF and INH monoresistance rates in Ghana were 0.8% (n=20) and 6.9% (n=172) respectively. S450L (n=35) and S531L (n=16) *rpoB* mutations (22.4%, n= 115), and V483G (n=6) and G332R (n=4) *rpo*C mutations (1.9%, n=10) predominantly conferred RIF resistance (Table S4; FIG. 4). S315T (n=364) mutation in *kat*G (74.5%, n=382) and G204D (n=32) mutation at the *inh*A promotor region (20.3%, n=104), was associated with INH resistance. However, Otchere *et al* (2016) detected novel mutations in *inh*A (G204D), in *ahp*Cpro (-88G/A and -142G/A), and in *ndh* (K32R) (61). The MDR rate in Ghana was 4.2% (n=104) (Tables S1-S3), which was lower than the finding reported from other African countries, but higher than those reported from Cameroon (3.7%), Tunisia (0.9%), and Morocco (2.6%).

#### Guinea Bissau

A resistance rate of 9% (n=9) was reported by Rabna *et al* (2015) in Guinea Bissau using 100 clinical isolates. S531L mutation in *rpo*B mediated RIF resistance (100%, n=10 isolates) (Tables S4; FIG. 4). INH resistance was not detected in this study. In 2016, the estimated incidence of MDR/RIF resistant-TB in Guinea Bissau was 11/100,000 (5).

#### Ivory Coast

Three research studies involving 261 clinical isolates in Ivory Coast yielded a resistance rate of 33% (n=86) (62–64) and an INH monoresistance rate of 6.5% (n=17). D516V (n=27) and S531L (n=12) mutations in *rpoB* conferred RIF resistance in 67.4% (n=58) of isolates whilst S315T mutation in *katG* (88.4%, n=76 isolates) and T8A/C (n=11/n=16) mutation in the *inhA* promoter (37.2%, n=32 isolates) conferred INH resistance (Tables S1-S3). The rate of MDR in Ivory Coast was 24.5% (n=64) (Figure 2d).

#### Nigeria

Dinic *et al* (2012) (65) and Senghore *et al* (2017) (38) characterized the mechanisms of first-and second-line anti-TB drug resistance in Nigeria, which had a resistance rate of 13.3% (n=65) from 488 clinical isolates. This result was similar to that of Tanzania (13.7%), but higher than those from Tunisia (3.2%), Malawi (6.4%), Rwanda (0.6%), and Guinea Bissau (9%). RIF monoresistance rate was 1.2%(n=6), mediated by S450L (n=32) and D435Y (n=10) mutations in *rpo*B (91.9%, n= 57) (Tables S1-S3). In 2016, the estimated incidence of MDR/RIF-resistant TB among notified pulmonary TB cases was 5200; and MDR/RIF-resistant TB among new and previously treated cases were 4.3% and 25%, respectively (5). From the same report, laboratory-confirmed cases of MDR/RIF resistant-TB cases were 1686 (5).

#### Sierra Leone

From 97 clinical isolates, a 51.5% (n=50) drug resistance rate and a 1% (n=1) RIF monoresistance rate were detected. S531L (n=8), H526R and H526Y (n=3) *rpo*B mutations (32%, n= 16 isolates) conferred RIF resistance, albeit certain *rpoB* mutations at codons 511, 516, and 533 conferred no RIF resistance (Tables S1-S3)(66). INH monoresistance rate, largely mediated by S315T (n=23) mutation in *kat*G and C15T (n=2) mutation at the *inh*A promoter, was 8.2% (n=8); high-level *katG-* and *inh*A promotor-mediated INH resistance was 56% (n=28 isolates) and 4% (n=2 isolates) respectively (Tables S4; FIG. 4) (66). STM monoresistance in Sierra Leone, mediated by K34R (n=11) and K88R (n=8) mutations in *rps*L, was 14.4% (n=14); *rps*L conferred STM resistance in 38% (n=19) of the isolates. In Congo, 19% of STM resistance was due to K43R *rps*L mutation (67). Similarly in Egypt, K43R and K88R formed 13.2% of *rps*L mutations (Tables S4; FIG. 4) (47).

### Central Africa: Cameroon, Chad, Gabon

#### Cameroon

Among 163 clinical isolates, the rate of anti-TB drugs resistance was 38.7% (n=63) (**Figure 2a**). S531L (71.4%), H526D (14.3%), and D516V (14.3%) *rpo*B mutations conferred RIF resistance in seven isolates (11.1%). S315T mutations (73% of isolates) in *katG* and mutations at -15 *inh*A promoter region conferred INH resistance in 30.2% (n=19) and 28.6% (n=18) isolates respectively (Tables S1-S4; FIG. 4) (68). The MDR rate in Cameroon was 3.7%(n=6)(68); although in 2016, the estimated incidence of MDR/RIF resistant-TB in the country was 6.8/100,000 (5).

#### Chad

Ba Diallo *et al* (2017) detected the emergence and clonal transmission of MDR-TB in Chad (24) to be 4.5% (n=14)(24); the estimated incidence of MDR/RIF-Resistant (RR) TB in 2016 was 5.3/100,000 population (5). However, the overall TB resistance rate in 311 isolates was 23.5% (n=73) while RIF monoresistance rate was 5.5% (n=17). S531L (n=15) and H526Y (n=5) *rpo*B mutations (23.3%, n= 17 isolates) mediated RIF resistance (24) while INH monoresistance, conferred by S315T1 (n=30) in *katG* (41.1%, n=30 isolates) and C15T(n=19) mutation at *inh*A promotor region (26%, n=19 isolates), was 13.5% (Tables S1-S4; FIG. 4).

#### Gabon

Alame-Emane and his colleges (2017), using GeneXpert remnants, reported a drug resistance rate of 16.9% (n=21) and an MDR rate of 14.5% (n=18)(69), albeit the estimated MDR/RR-TB incidence in Gabon in 2016 was 20/100,000 (5). S531L and D516Y (n=6) *rpo*B mutations conferred 100% RIF resistance (n=21) while *kat*G (76.2%, n=16) and *inh*A (81%, n=17) mutations conferred INH resistance (Tables S1-S4; FIG. 4).

### East Africa: Ethiopia, Rwanda, Tanzania, Zambia, Uganda

#### Ethiopia

Next to South Africa, Ethiopia reported many studies (12 studies; 2,779 clinical isolates) that characterised TB drug resistance mechanisms, totalling a resistance rate of 35.7% (n=991) and an MDR rate of 23.9% (n=665). RIF monoresistance rate in Ethiopia was 3.1% (n=84), mediated by S**53**1L (n=485) and H526Y (n=55) mutations in *rpo*B (62.6%, n=612). INH monoresistance rate in Ethiopia was 3% (n=84), conferred by S**315**T(n=633) in *kat*G (79.1%, n =784) and C**15**T (n=22) mutation at *inh*A promotor region (4.5%, n =45). The highest MDR rate was reported by Abate *et al* (2014), which was 58% (n=427) (48). However, the 2016 estimated incidence of MDR/RR-TB was 5.7/100,000, and MDR/RR-TB case among new and previously treated cases were 2.7% and 14%, respectively (5). Similar findings were reported from Ivory Coast (24.5%) (63), although its higher than that of other African countries (Tables S1-S4; FIG. 4).

#### Rwanda

Umubyeyi *et al* (2007) determined the rate and mechanisms of fluoroquinolone resistance among 701 isolates in Rwanda to be 0.6% (n=4) and *gyr*A (100%, n=4)-mediated, respectively (60); T80A (n=3) and N94A (n=1) *gyr*A mutations mediated ofloxacin (OFL) resistance. OFL resistance rate in Mozambique was however 1.9% (n=6) (58).

#### Tanzania

Three studies, comprising 226 clinical isolates, yielded a resistance rate of 13.7% (n=31) in Tanzania while the MDR rate was 6.2% (n=14). The highest MDR rate was reported in 2013, i.e. 59.1% (n=13/22), albeit a significant reduction of 1.4% (n=1/74) was reported by Kidenya *et al* (2018) (70). Estimated MDR/RR-TB rate in new and previously treated cases were 1.3 % and 6.2%, respectively in 2016 (5). RIF, INH, EMB and STM monoresistance rates were 0.9% (n=2), 3.1% (n=7), 0.9% (n=2), and 1.8% (n=4) respectively. STM monoresistance however dropped from 2.3% (n=3) in 2007 (71) to 1.4% (n=1) in 2018 (70) (Tables S1-S3). S**531**L(n=4) mutation in *rpo*B (54.8%, n=17 isolates), *kat*G (58.1%, n=18 isolates) and *inh*A (9.3%, n=3 isolates) mutations, M306V/I and E378A (n=2) mutations in *ebm*B (29%, n=9 isolates), as well as K43R (n=2) mutation in *rps*L (6.5%, n=2 isolates) respectively mediated RIF, INH, EMB and STM resistance (Tables S4; FIG. 4).

#### Zambia

The resistance rate of 123 isolates in Zambia was 26% (n=32), with a PZA monoresistance rate of 3.3% (n=4), which was higher than previous reports from other African countries. S65S (n=17) and L35R (n=7) mutations in *pnc*A (84.4%, n=27) conferred resistance to PZA (Table S1-S4; Fig. 4).

#### Uganda

A resistance rate of 31.8% (n=215) and an MDR rate of 18.3% (n=124) was obtained from four studies involving 676 clinical isolates. RIF and INH monoresistance rates were respectively 2.7% (n=18) and 9.3% (n=63) (Tables S1-S3). S531L (n=46) and S450L (n=10) in *rpo*B (50.2%, n=108) as well as *kat*G 60.5% (n=130) and *inh*A promotor region (4.7%, n=10) mutations were respectively implicated in RIF and INH resistance (Fig. 4; Table S4). A higher INH monoresistance rate of 60% (n=54) was reported in 2016 (32) while an MDR rate of 82.4% (n=42) was detected in 2013 (72). In 2016, the estimated incidence of MDR/RR-TB in Uganda was 4.7/100,000; however, a 2011 national survey reported that MDR/RR-TB cases among new and previously treated cases were 1.6% and 12%, respectively (5).

### SOUTHERN AFRICA: Botswana, Malawi, Mozambique, Swaziland, South Africa, Zimbabwe

#### Botswana

The overall resistance and MDR rates of 260 clinical isolates in Botswana were respectively 7.7% (n=20) and 1.5% (n=4), with an INH and RIF monoresistance rate of 4.2% (n=11) and 1.9% (n=5) respectively. Resistance to RIF and INH were mediated by *rpo*B (95%, n=19 isolates) and *kat*G (65%, n=13 isolates) gene mutations (Tables S1-S4; Fig. 4)

#### Malawi

In Malawi, the overall TB resistance rate among 995 clinical isolates was 6.4% (n=64). H526Y (n=11) and S531L (n=10) mutations in *rpo*B 57.8% (n=37) mediated RIF resistance (Tables S1-S4; Fig 4).

#### Mozambique

Two studies in Mozambique that comprised 323 *M. tb* isolates, resulted in a resistance rate of 32.8% (n =93) and an MDR rate of 15.5% (n=50). The estimated TB cases with MDR/RR-TB in new and previously treated cases were 3.7% and 20%, respectively in 2016(5) (Tables S1-S3). The monoresistance rates of RIF, INH and EMB were 0.6% (n=2), 8% (n=26), and 2.8% (n=9) respectively. S531L (n=10) and H526Y (n=6) mutation in *rpo*B 24.5% (n=26), S**315**T (n=64) mutation in *katG* (69.8%, n=74) and C15T (n=9) mutation in *inh*A (10.4%, n=11) promotor region, and M306I/V (n=3/n=2) mutation in *emb*B (11.3%, n=12) conferred RIF, INH and EMB resistance. Furthermore, A90V (n=7) and D94G (n=2) mutations in *gyr*A 9.4% (n=10) mediated OFL (Fluoroquinolones) resistance (Fig. 4; Table S4)

#### Swaziland

Andre *et al* (2017) and Sanchez-Padilla *et al* (2015) characterised drug-resistant *M. tb* with an Xpert MTB/RIF and a novel rapid PCR, which detected an I491F mutation in *rpo*B (73, 74). Notably, both the resistance and MDR rates among 203 isolates was 100% (n=203).

#### South Africa

The overall TB resistance rate in South Africa was determined to be 16.8% (n=8197) among 48 760 isolates, which was lower than that of Sudan (67.9%), Zimbabwe (100%), and Burkina Faso (57.1%). The MDR rate was 4.5% (n=2189) and was lower than that of Sudan (34.6%), Ethiopia (23.9%), Ivory Coast (24.5%), and Burkina Faso (46%). The 2016 MDR/RR-TB incidence was 34/100,000 population and the estimated rate of TB cases with MDR/RR-TB among new and previously treated cases were 3.4% and 7.1%, respectively (5). In the same report, the estimated MDR/RR-TB cases among notified pulmonary TB cases was 8200, whilst laboratory confirmed MDR/RIF resistant-TB cases was 19073 (5). Out of 33 studies that determined the resistance mechanisms of first- and second-line drugs, 12 reported on the XDR rate, which was found to be 1.6% (n=781). Laboratory confirmed XDR-TB cases in South Africa in 2016 was 967 (5).

Monoresistance rates of RIF, INH, EMB, STM, CAP, KAN and OFL were 0.01%(n=4), 0.3% (n=163), 4.2% (n=2049), 0.01(n=7), 0.1% (n=31), 0.04% (n=20) and 0.01% (n=3). S531L(n=478), D616V (n=149), and 516GTC (n=167) mutations in *rpo*B (25%, n=2053), A542A (n=80) and V483G (n=48) mutations in *rpo*C (2.4%, n=193) and *rpo*A 0.05% (n=4) mutations respectively conferred high-level, low-level and low-level RIF resistance (Fig. 4; Tables S1-S4).

The predominant mechanisms conferring resistance to INH were S315T (n=672) in *kat*G (21.9%, n=1798) and C15T (n=294) at *inh*A promotor region (9.2%, n=758) whilst M306I mutation (n=472) in *emb*B (7%, n= 573) mediated EMB resistance. In addition, A1401G (n=639) and A513C (n=398) mutations in *rrs* (15.1%, n=1241), K43R (n=10) mutation in *rps*L (0.1%, n=12) and *eis* mutation (n=13) mediated resistance to STM and CAP; *rrs* confers mono and cross resistance to CAP (60.5%, n=75). A high frequency mutation was observed at the rrs1400 (n=26) and rrs900(n=22) loci of *rrs* (75). As well, D94G (n=218) and A90V (n=168) mutation in *gyr*A 8.9% (n=728) and mutations in *gyr*B (0.7%, n=61) mediated resistance to OFL (Figure 4; Table S4).

#### Zimbabwe

Takawira *et al* (2017) characterized *rpo*B and *kat*G genes in MDR-TB isolates (100%) to respectively determine RIF and INH resistance mechanisms (18). RIF resistance was mainly mediated by *rpo*B (93.5%, n=86) mutations such as S531L and novel ones such as G507S, T508A, L511V, del513-526, P520P, L524L, R528H, R529Q and S531F; novel mutations such as I572P/F, E562Q, P564S, and Q490Y were outside the RIF resistance determining regions (Tables S1-S3). Inconsistencies between genotypic and phenotypic assay results for both *kat*G and *rpo*B genes were observed (18).

### Molecular epidemiology: Lineage and family distribution

All the seven lineages of *M. tb* species and *M. bovis* were found in the included studies with varying distribution frequencies per country (Figures 6 and S1). The Euro-American Lineage (Lineage 4) had the largest frequency distribution among all the lineages in Africa, followed by Beijing (Lineage 2), Central Asian (Lineage 3), West African I (Lineage 5) and II (Lineage 6), Indo-Oceanic (Lineage 1), Ethiopian (Lineage 7) and *M. bovis* (Lineage 8) lineages. The frequency distribution of the sub-lineages among these eight lineages are shown in Figure S1, and these varied per country. For instance, most of the Beijing strains were reported from South Africa whilst the T and LAM sub-lineages were very common in Northern, Eastern and Southern African countries; specifically, the T lineages were very dominant in most of the countries (Fig. 6). The H family was also commonly found in Northern Africa. As expected, the CAS strains were very common in Central and Eastern Africa, Madagascar, and Malawi, whilst the West African I and II strains, the Cameroon strains, and Sierra Leone strains were mainly found in West African countries such as Nigeria, Ghana, Cameroon, Chad, Senegal, and Sierra Leone. Furthermore, the Congo and Uganda strains were more prevalent in Central and Eastern Africa whilst EAI lineages were reported from mainly Guinea, Guinea Bissau, Eastern and Southern African countries (Fig. 6 and S1; Supplementary dataset 2).

### Phylogenomics: evolutionary genomic epidemiology

The relationship between deposited resistant and susceptible *M. tb* isolates at GenBank/PATRIC is shown in Figure 7, with Figures 7a and 7b being phylogenomic trees of both resistant and susceptible strains while Figures 7c and 7d represent only resistant strains. The lineages and sublineages of the individual strains were not included in their metadata and could thus not be included in the trees. Most of the isolates were from South Africa, Swaziland, Mali, and Uganda, and these are shown as unique colours on the trees. Most of the resistant isolates were from South Africa. Notably, there was relatively minor trans-boundary transmission of closely related strains/clones while most of the isolates were localized within countries.

**Figure 7 (A-D):**
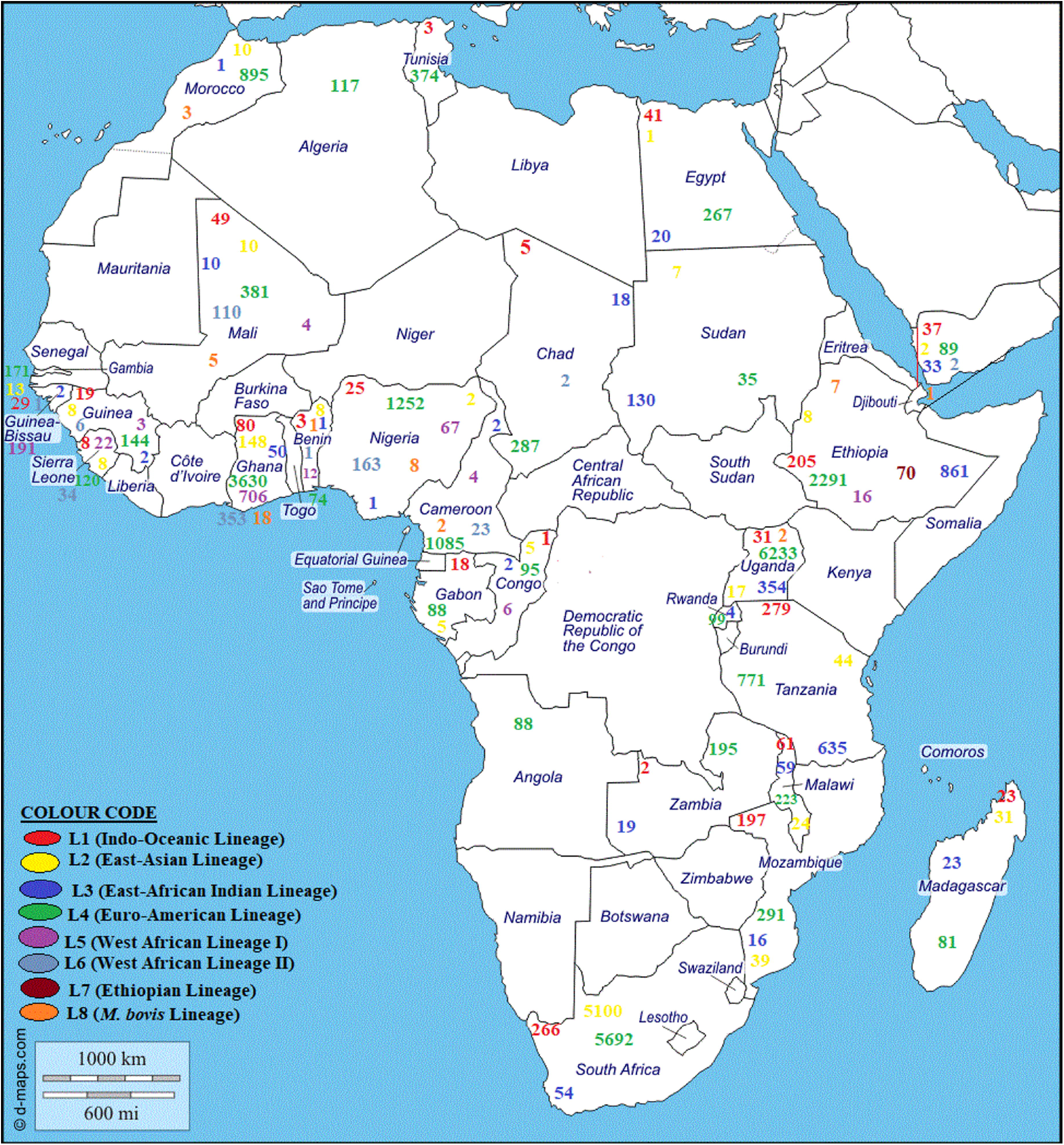
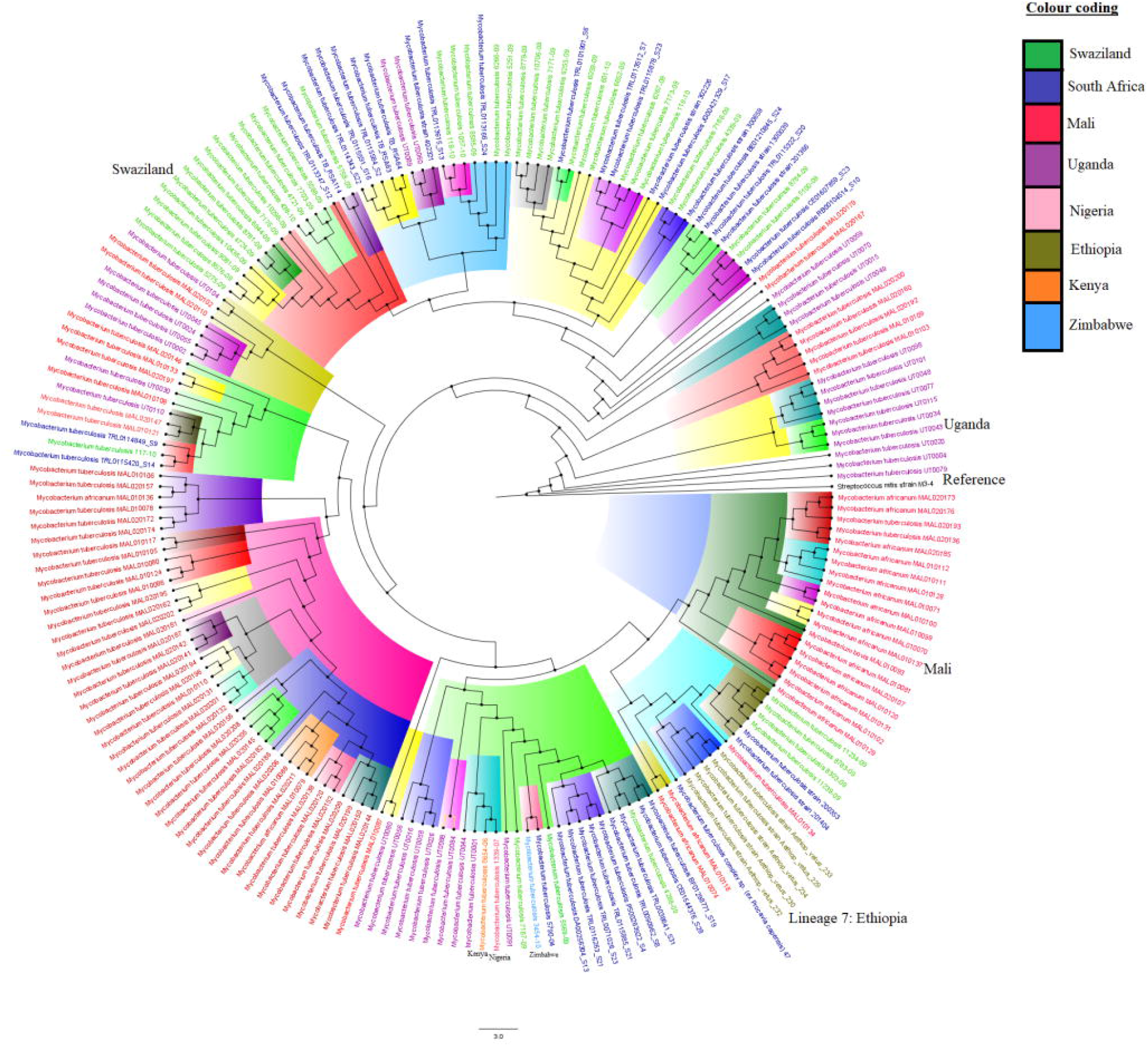
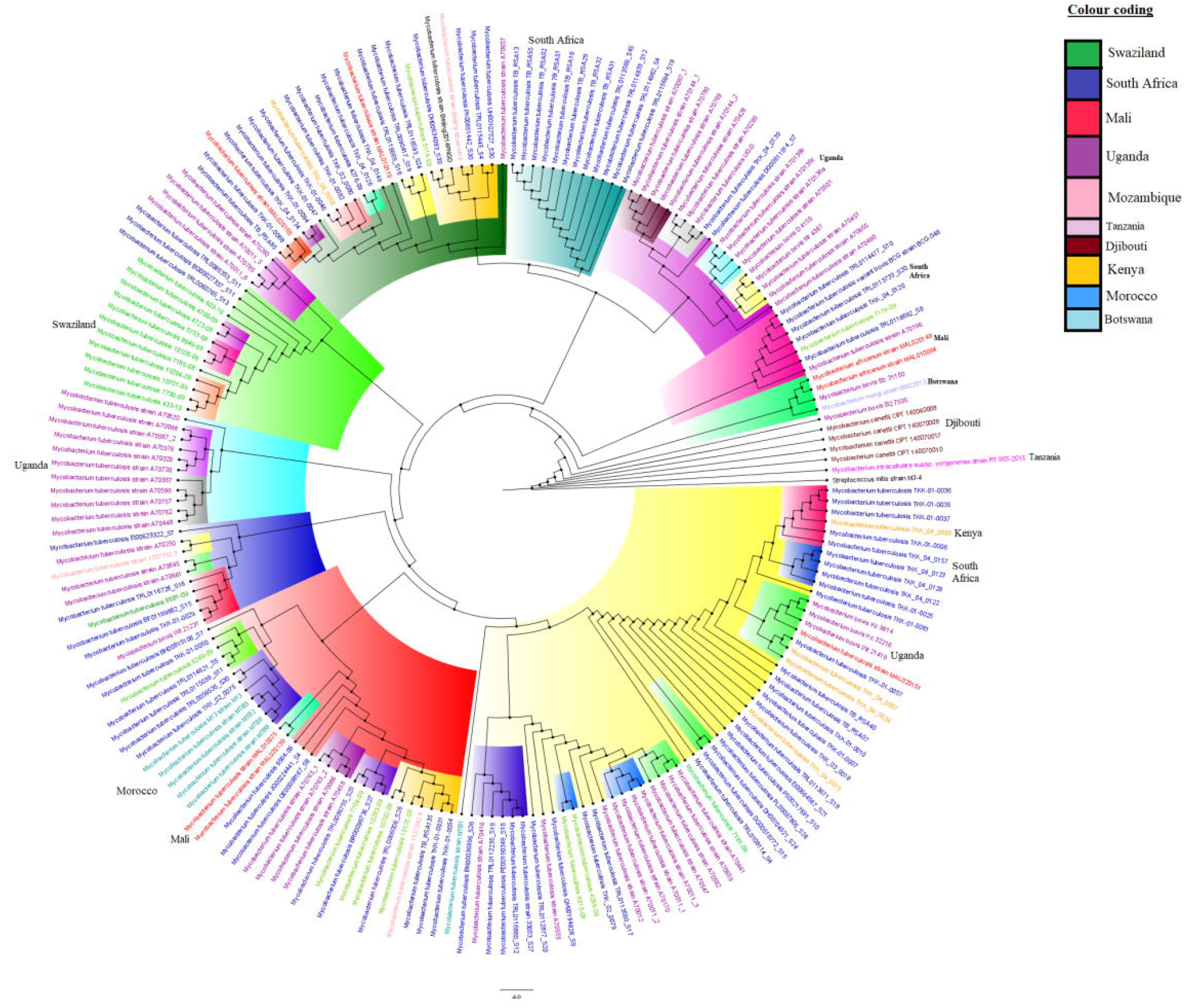
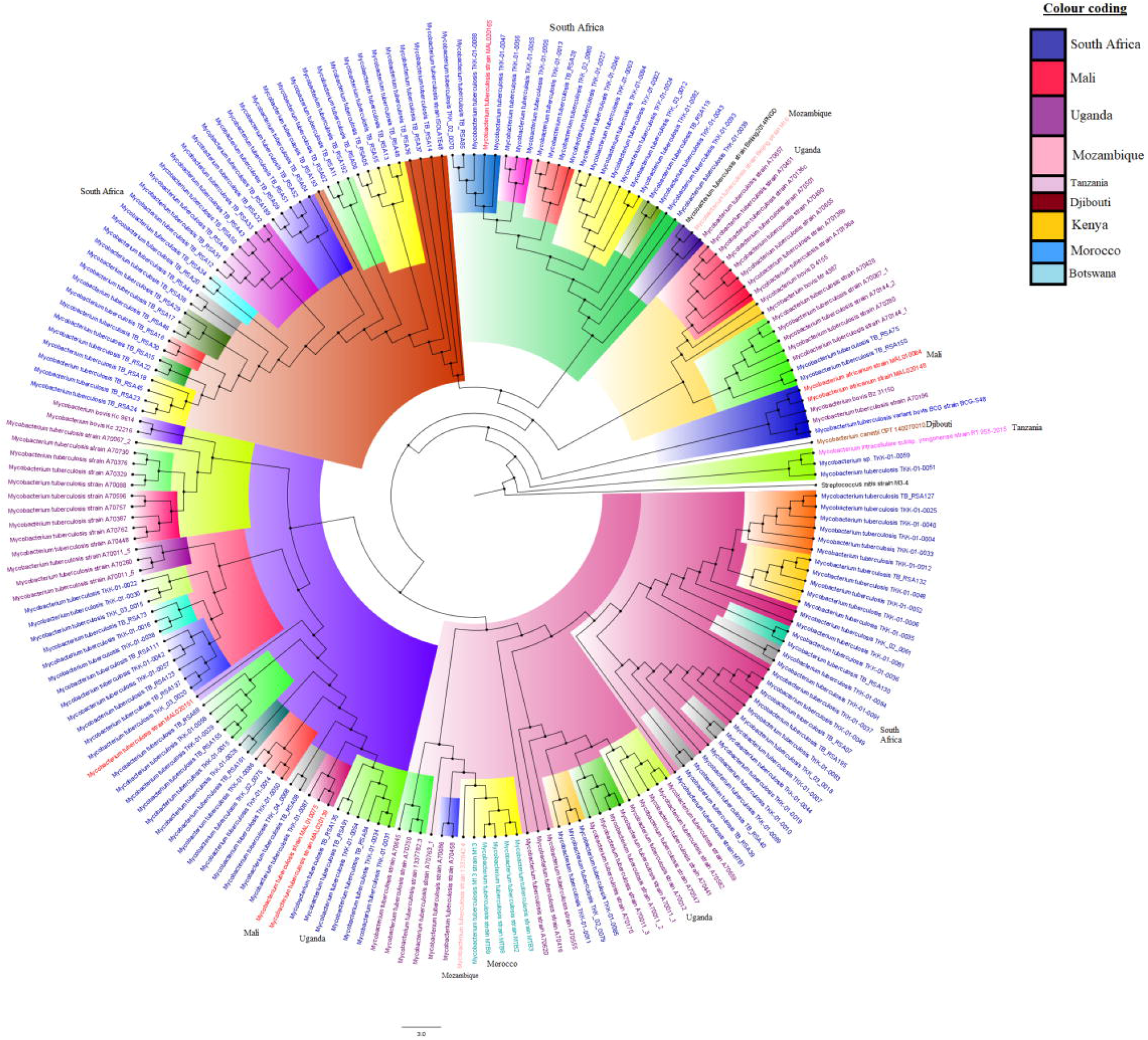
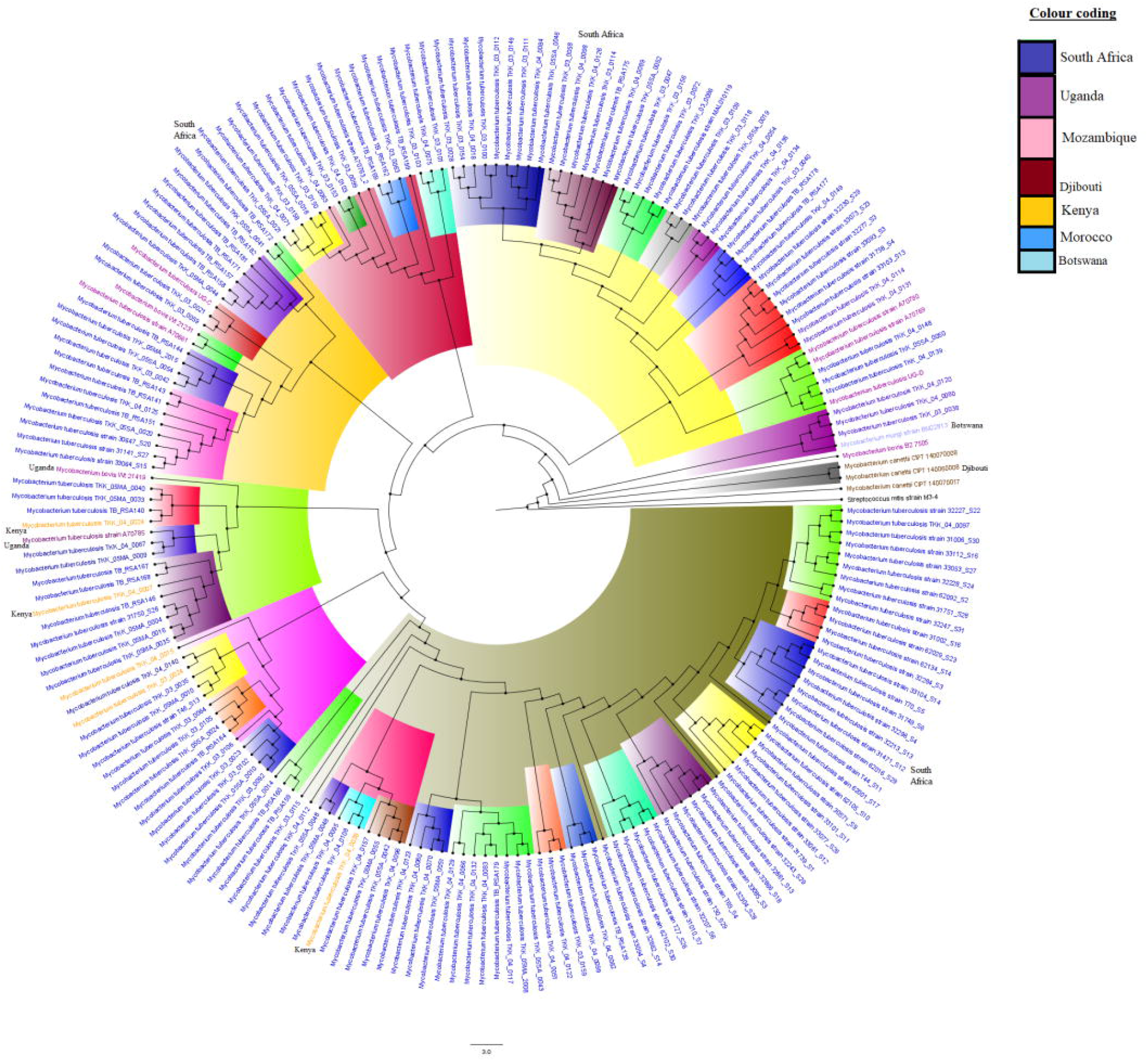
Phylogenomic relationship between *M. tuberculosis* isolates of African origin deposited at GenBank and PATRIC. Figures 7A and 7B comprise of both resistant and susceptible strains while Figures 7C and 7D comprise of only resistant strains. Strains from the same countries are given the same colour labels while those from the same clade and sub-clade are highlighted with the same colour to show their closer relationships. As can be seen, there was more intra-country dissemination and evolution than inter-country spread.

### Diagnostics

Phenotypically, the BACTEC MGIT 960 system and the agar proportion methods were commonly used to diagnose *M. tb* and determine antibiotic sensitivity from clinical specimens. The methods and instruments used differed from country to country (Tables 2, S3 & S5). Molecular tests such as PCR and amplicon sequencing, Hain’s line probe assays (GenoType®MTBDR), GeneXpertMTB/RIF and currently, WGS, were commonly used in 122 studies to both identify and determine drug resistance in *M. tb*. More studies from South Africa (n=33) and Ethiopia (n=13) used molecular methods to both detect *M. tb* and ascertain its resistance profile (Tables S3 & S5).

## 4. Discussion

The epidemiology of antitubercular drug resistance in Africa, as shown in this review, is phylogenetically limited to countries than across countries, and were mediated by non-lineage-specific mutations. Notably, INH and RIF, which are the most important and main first-line anti-TB drugs, had the most resistance prevalence in many African countries. The high proportion of INH resistance could be due to the relative ease and alacrity with which resistance mutations developed to this antibiotic. The genes in which mutations usually occur to confer resistance to first-and second-line anti-TB antibiotics include: *rpo*B, *rpo*A, and *rpo*C (RIF); *kat*G and *inh*A (INH); *pnc*A (PZA); *emb*CAB (EMB); *rps*L, *rrs*, and *gid*B (STM); *rrs* and *eis* (KAN/AMK); *rrs* and *tly*A (CAP); *gyr*A and *gyr*B (Fluoroquinolones); *eth*A, *eth*R, and *inh*A (ETM); *thy*A and *rib*D (Para-aminosalicylic-acid); *alr, ddl*, and *cyc*A (Cycloserine); *atp*E (Bedaquiline); *rrl* and *rpl*C (Linezolid) (2, 27, 76). RIF is a broad-spectrum drug that interferes with the synthesis and elongation of mRNA by binding to the *β-*subunit of RNA polymerase, an enzyme encoded by the *rpo*B gene in *M. tb*. Mutations occurring in the 81bp hypervariable region of the *rpo*B gene reduces the binding affinity of RIF to RNA polymerase, leading to RIF resistance in 96% of RIF-resistant isolates (29). RIF resistance-conferring mutations in the *rpo*B gene mainly occurs between codon 507 and 533, which is called the RIF-resistance-determining region (RR-DR) (48, 49). In most cases, mutations that occurred at codon 531 and 526 results in RIF resistance (48). Abbadi *et al* (2009) observed that 76% of resistance-conferring mutations in the *rpo*B gene in Egypt was due to S**351**L (47). Abate *et al* (2014) also revealed that 68.7% of RIF-resistant strains in Ethiopia had mutations at codon 531 (S**531**L) (48). Moreover, *rpo*B mutations at codons 531 and 526 have been reported by several studies in different African countries to confer RIF resistance (36, 47, 48, 64, 77–81). As well, *rpo*B mutations at codon 516 has been reported by other studies to confer RIF resistance (Tables S1-S4).

INH is a prodrug that needs activation by a catalase enzyme encoded by the *kat*G gene. Mutations in the *kat*G gene, most commonly occurring at codon 315, lower catalase-peroxidase activity and result in high-level INH resistance. Feuerriegel *et al*. (2012) (66) and Abate *et al* (2014) (48) respectively found that 72% and 93% of *kat*G gene mutations occurred at codon 315 (S**315**T) in Sierra Leone and Ethiopia. These have been corroborated by several studies from different African countries, which also found mutations at codon 315 in *kat*G as mediating high-level INH-resistance (Figures 3–6; Tables S1-S4). Mutations at the promotor region of *inh*A also induce low-level INH resistance (Figures 3–6; Tables S1-S4)(29). The *inh*A mutations also mediate resistance to another second-line drug, ETM, due to the similar molecular structure of the two drugs (29). Activated INH prevents the synthesis of mycolic acids by inhibiting NADH-dependent enoyl-ACP reductase, which is encoded by *inh*A (29). N’Guessan *et al*. (2016) showed that 45% of *inh*A mutations contributed to INH resistance in Ivory Coast(63).

The high proportion of INH resistance could be due to the easy propensity for resistance mutation to be developed in *katG* and *inhA*. Ba Diallo *et al*. (2017) in Chad, from their study on the emergence and clonal transmission of MDR-TB, found 13.5% INH-monoresistance (24) while N’Guessan *et al* (2008) in Côte d’Ivoire reported 8.7% INH-monoresistance (64); three previous studies in Egypt reported 7%, 4%, and 12.5% INH-monoresistance rate (2, 47, 54). Considerable proportion of INH mono-resistance rate i.e., 1.7%-11.6%, was also reported from Ethiopia (12, 78, 83, 84), and 7.4%-10.9% in Ghana (19, 61, 85), with INH monoresistance in Ghana increasing from 2015 to 2018: in 2015, it was 8.2% (n=43/525) (19), in 2016 it was 7.4% (n=111) (61), and in 2018, it was 10.9% (n=18) (85). However, the highest INH-monoresistance rate, i.e. 13.5 % (42 INH mono-resistant isolates), was in Chad (24) (**Figure 2b**).

EMB, a first-line TB drug, interferes with the biosynthesis of cell wall arabinogalactan (66). Several studies reported that 50% of EMB resistance is mainly due to mutation in *emb*B gene at codon 306 (66, 81). In agreement to this, other previous studies have shown that common *emb*B gene mutations conferring EMB resistance occurred at codon 306 (32, 35, 45, 72, 86, 87). Feuerriegel *et al* (2012) (66) and Dookie *et al* (2016) (88) revealed that 46.7% and 86% of EMB resistance was due to *emb*B gene mutation(s) at codon 306 in Sierra Leone and South Africa respectively; in the study by Dookie *et al* (2016), 90.7% of isolates with EMB resistance had mutations in the *emb*B gene (88). In addition, other common mutations in *emb*B, besides those at codon 306, confer EMB resistance in different African countries (32, 35, 45, 72, 86, 87). EMB monoresistance in Sudan was 1.3% (n=3) (11), but the mechanisms of resistance (gene mutation) was not determined, which could be due to the diagnostic they used. This result was similar to that from Egypt, which was 1.9% (2, 47); but less than that of Mozambique (2.8%, n=9) (58) and higher than that of South Africa (0.01%, n=4) (89).

STM is an aminocyclitol glycoside antibiotic, which hinders the initiation of translation by binding to the 16S rRNA. STM resistance is mostly mediated by mutations in the *rrs* or *rps*L gene, which alters the STM-binding region to prevent STM binding. Mutations in *rrs* and *rps*L genes have been identified in more than 50% of the STM-resistant isolates, with the most common *rps*L mutation being at codon K**43**R and K**88**R (29). Aubry *et al* (2014) reported that 19% of STM-resistance was due to K**43**R mutation in *rps*L in Congo (67). A similar finding was reported by Abbadi *et al* (2009) in Egypt, where 13.2% of *rps*L mutations occurred at codon K**43**R and K88R (47). Similar findings of *rps*L mutation at K**43**R and K**88**R has been reported by other studies (32, 36, 38, 66). Furthermore, mutation(s) in the *gid*B gene, which encodes a conserved 7-methylguanosine methyl transferase specific for the 16S rRNA, can mediate low-level STM resistance. Tekwu *et al*. (2014) revealed that 11.1% (n=7) of STM-resistance was due to *gid*B mutation at different hotspot regions in Cameroon (68). One study from Sierra Leone revealed that 50% of STM-resistance was due to *gid*B mutation (66).

The STM mono-resistance rate in Africa was found to be 0.1% (57 STM mono-resistant isolates), being lower than that reported in individual countries: 14.4% (66) in Sierra Leone, 10% in Sudan and 7.9% in Egypt. The highest EMB monoresistance was reported from Mozambique i.e. 2.8% (n=9 isolates). Monoresistance to all the three second-line injectable drugs was reported from South Africa: CAP (0.05%, n=31 isolates), kanamycin (KAN) (0.03%, n=20 isolates) (Table S1-S3).

Feuerriegel *et al* (2012) conducted molecular sequencing analysis of first-line anti-TB drugs to characterize the mechanisms of resistance. The study identified different amino acid/codon changes in drug sensitive strains as well (66), implying that the presence of different amino acid/codon change at the hotspot region of certain genes does not corroborate a drug-resistant phenotype.

### MDR & XDR rates

MDR rates in the various countries are substantial and worrying, given the higher mortalities associated with MDR phenotypes. Besides South Africa, Ethiopia, and Ghana, where a substantial number of studies involving numerous isolates were used, the MDR rates in most of the studies undertaken in the other countries involved relatively few strains. Nevertheless, countries with lesser sample numbers but higher MDR rates requires urgent attention. For instance, Swaziland and Zimbabwe had very high MDR rates although their sample sizes were smaller. The dearth of comprehensive studies to identify MDR strains in Africa should be urgently addressed to aid policy decisions with regards to TB interventions. Furthermore, XDR rates were obtained only from South Africa as the studies from other countries did not provide such data. This implies that other African countries had limitations to detect XDR-TB, due to either financial constraints or unavailability of molecular diagnostic assays for detecting and characterizing Pre-XDR and XDR-TB. This confirms the need to increase *M. tb* resistance epidemiology studies in Africa to inform evidence-based decisions.

The MDR rates obtained in this study either varied or agreed with those from the national survey reports of the respective countries. The MDR rate in morocco was 2.6% (n=26) and was comparable with the 2016 estimated incidence rate of MDR/RR-TB cases, which was 1.8/100,000 (5); the 2016 national survey reported that the MDR/RR-TB cases among previously treated cases was 11% (5). However, the 2016 national Sudan survey report showed that the MDR/RR-TB in new and previously treated case were 2.9% and 13%, respectively; the incidence rate of MDR/RR-TB was 3.1/100,000 (5). This result was higher than the report from Morocco (2.6%) (23), Egypt (12%) (47), and Ethiopia (23.9%) (48). The MDR rate in Tunisia i.e. 0.9% (n=12) (90) (Figure 2D), tallied with the country’s 2016 estimated incidence of MDR/RR-TB, which was 0.4/100,000; MDR/RR-TB cases among new and previously treated cases were 0.93 % and 4.2%, respectively (5). In 2016, the estimated incidence of MDR/RR-TB in Ghana was 3/100,000; however, the 2017 national survey reported that MDR/RR-TB case among new and previously treated cases were 1.5% and 7%, respectively (5).

In neighboring Ivory Coast, an MDR rate of 87.3% (n=55) (63), which was virtually similar to the 2017 national survey report, was reported. In 2016, the estimated incidence of MDR/RR-TB in Ivory Coast was 8.9/100,000 whilst the 2017 national survey on MDR/RR-TB case among new and previously treated cases were 4.6% and 22%, respectively (5). The MDR rate in Egypt was comparable to the 2011 national survey report, which had 14% MDR/RR in new TB cases, but lower than the 2016 national survey report for MDR/RR cases (20%) among previously treated cases (5).

### Molecular epidemiology

Molecular genotyping is a significant tool for tracing genotype/strain distribution in different geographical settings. These techniques can predict transmission rates and identify dominant strains in limited settings, strains that stand a chance of spreading and those related to outbreaks (42), severe disease (41), and drug resistance. Isolates with a very close phylogenomic relationship are more likely to have evolved from the same ancestor at some point and their presence in the same patient may suggest within-host evolution or exposure to multiple patients who were infected from the same or closely related strains. Such closely related strains provide invaluable data on disease transmission epidemiology (91). Spoligotyping, MIRU-VNTR and/or whole genome sequencing techniques used in the included studies reported diverse *M. tb* genotypes/lineages across Africa; out of the 41,520 isolates characterized/genotyped, only 99.7% (41,403 genotypes) were identified and found under the international database.

Notably, both the phylogenomics and the lineage/family distribution per country support localized or regional distribution of specific strains, although the T and LAM families under lineage 4 were more extensively distributed throughout Africa. The extensive distribution of lineage 4 and its specific sub-lineages such as T throughout Africa is worthy of attention, particularly as the reason for this is unknown. The country- and region-specific prevalence of the lineages suggests limited intercountry spread of *M. tb* in Africa as there were more intra-country dissemination of *M. tb* strains, with KZN strains being mainly found in the KwaZulu-Natal Province of South Africa. This could suggest a limited intercountry migration in Africa. Moreover, the presence of diverse lineages in South Africa could be also explained by the immigration of several Africans, Asians and Europeans into South Africa.

We could not establish a relationship between individual strains/lineages and specific mutations, contrary to what has been reported. For instance, atypical Beijing lineages in Africa were shown to be associated with specific amino acid changes in genes including *rpo*B (D516V), *kat*G (S315T), *inh*A (G17T), *emb*B (M306I), *rrs* (A513C, A1401G) (35, 45, 86). Muller *et al*, (2013) mentioned that R86 strains have high degree of clustering for drug resistance mutation patterns that affects more than 40% of all MDR patients, including mutations associated with resistance to first and second-line antibiotics (35). However, meta-analyses of the collated data shows that these mutations are also found in other strains and lineages in almost all countries. The presence of similar mutations in different lineages and countries suggests that such mutations are not lineage or country specific (Table S4 & S6). We thus believe that the limited view provided by the relatively small sample sizes used in individual studies per country could not provide a wholistic overview of resistance mutation distribution per lineage, leading to a skewed conclusion.

In cases where lineages or families in certain countries and studies have been shown to be more resistant to certain antibiotics, caution should be exercised in generalizing such results as the local TB management culture, diagnostics used and endemic/circulating drug-resistant strains could be determining factors. Even when lineages and families were found to be more resistant to certain antibiotics, the underlying resistance mechanisms differed between studies and/or lineages/families. Furthermore, where such lineage-specific mutation observations have been made in one country, it has not been found to be the case in all other countries, limiting its widespread application (Tables S1-S6). Some few examples below shall suffice.

Ba Diallo *et al*., in Chad, identified that SIT61(Cameroon family) had higher proportion of resistance to RIF, INH and PZA, being respectively mediated by mutations in *rpo*B (S531L), *kat*G (S315T1) and *pnc*A (Del143R) (24). In agreement to this, Otchere *et al* (2016) in Ghana reported that Lineage-4 (Cameroon family) had higher rates of RIF and INH resistance, also mediated by mutations in *rpo*B (S450L), *rpo*C (G332R), *kat*G (S315T) and *inh*A (G204D), respectively (61). In Ethiopia, the Haarlem sub-lineage was more resistant to INH, with a common gene mutation at *kat*G (S315T) (83), whilst the CAS-Delhi sub-lineage was more resistant to RIF through mutations at *rpo*B (Codon 447-452) (92). Furthermore, Feuerriegel *et al*.(2012) showed that drug-resistant LAM strains harboured various resistance mutations in *rpo*B (D516Y and S531L), *kat*G (S315T and Q471R), *inh*A (C15T), *emb*B (M306I and W332R), *rps*L (K43R), and *gid*B (G69D, G92D, L16R, A161A and Q127Stop) (66). As well, Bwalya *et al*., (2018) observed that CAS1-KILI expressed resistance to PZA, mediated by mutations at codons S65S and D8N while LAM11-ZWE had mutations at codons Q10R, D80N, G97A, W119C, R154G and A171V (50)(Table S4 & S6). The presence of the above-stated mutations in other lineages and countries is evident in Tables S1-S4 & S6, making it difficult to conclude on a country- or lineage-specific association of resistance mutations.

### Diagnostics used for detecting TB and determining TB resistance in Africa

In several resource-limited African countries, diagnosing drug-resistant TB is more challenging due to inadequate bacteriological laboratory facilities and/or skill (20, 82). Molecular diagnostic methods have an advantage over phenotypic methods particularly due to their short turnaround time and accurate diagnostic result (20). A major concern with molecular tests is their ability to detect *M. tb* DNA even in latent or cured TB patients, resulting in false positives. This is because TB DNA ca persist in patients long after treatment. is the Currently, different molecular diagnostic methods are available to test mechanisms of drug resistance (gene mutations) to a particular antibiotic (20, 26). Essentially, they all work through a DNA amplification technique and can be classified into real-time polymerase chain reaction (RT-PCR) assays, solid phase hybridization assays and sequencing technologies (20). The recent advances and applications of WGS is however revolutionizing the diagnosis of TB and drug resistance owing to its higher sensitivity, specificity, typing resolution and comprehensiveness, although its use in

Africa is still limited due to its higher cost and advanced skill required (Tables 2, S1-S3) (93). Unlike culture-based tests, molecular methods do not need viable cells to diagnose TB and TB resistance, which can also lead to false positives as TB DNA can remain in the host long after its death (94).

#### Line Probe Assay

The line probe assays (LPAs), GenoType^®^MTBDR*plus* and GenoType^®^MTBD*Rsl* (Hain Lifescience, Nehren, Germany) are the most commonly performed molecular diagnostic tests to detect and characterise mechanisms of drug resistance in Africa (Tables 2, S1-S3, S5). GenoType^®^MTBDR*plus* can detect MDR-TB i.e., resistance to RIF and INH by detecting *rpo*B, *kat*G and *inh*A promoter region mutations whilst GenoType^®^MTBD*Rsl* can detect XDR-TB by identifying both *M. tb* and its resistance (*gyr*AB mutations) to fluoroquinolones (e.g. ofloxacin and moxifloxacin) and second-line injectable drugs i.e., *rrs* and *emb*B (17, 58). LPAs use immobilized probes on a solid support to detect specific *M. tb* DNA targets and resistance-conferring mutations from smear-positive sputum samples or culture isolates, after extraction and amplification of the DNA (79, 82); the probes are attached to a nitrocellulose strip (77).

The sensitivity and specificity of MTBDR*plus* (version 2) in detecting RIF and INH from culture were 100% and 87.9% respectively for RIF, and 100% and 94.4% respectively for INH (79). The same study evaluated the sensitivity and specificity of MTBDR*sl* (version 1 and 2) in detecting EMB and other second-line drugs from cultured isolates and reported the following: EMB (60% and 89.2%), OFL (100% and 91.4%), KAN (100% and 97.7%); agreements between the LPAs and MGIT were very high for INH (k=0.93), good for RIF, OFL, and KAN (k=0.6–0.7) and moderate for EMB (k=0.5) (79). Another study showed the sensitivity and specificity of MTBDR*plus* to detect RIF, INH and MDR to be respectively 98.9% and 99.4% for RIF, 94.2% and 99.7% for INH, and 98.8% and 100% for MDR compared with conventional results (26). Similar studies by Bedawi *et al*.(2016) revealed that the sensitivity and specificity of MTBDR*plus* for RIF and INH resistance were 80% and 99.6%, and 82.7% and 99.6% respectively while the MDR results were 75% and 100% respectively (12).

MTBDR*plus* has several limitations in its application. For example, the amino acid sequence cannot be screened; hence, silent mutations(s) in the probe region can result in the absence of one of the wild type bands. Moreover, this method only screens the gene mutation in the locus of interest (target region) in the *rpoB, kat*G and *inh*A gene regions (33). Resistance due to gene mutation outside of the gene locus of interest will not be detected. Finally, MTBDR*plus* may not be used for monitoring the progression or success of treatment of patients with antimycobacterial therapy.

#### Xpert MTB/RIF assay

The GeneXpertMTB/RIF assay, which was endorsed by WHO in 2010 for the accurate laboratory diagnosis of TB and drug resistance as well as to increase case detection rates in resource-limited countries, is another commonly used diagnostic method in Africa (Tables 2 & S5). It is a real-time PCR-based technique used for detecting *M. tb* and RIF resistance using a single-use cartridge with a turnaround time of 2 hours (1, 94). The detection of *M. tb* and its RIF resistance can be done in an integrated and closed system, and sample processing is automated (95). Unlike the LPA, GeneXpert can only detect TB and RIF resistance and has low sensitivity to smear-negative samples. Helb *et al*. (2010) observed an overall sensitivity of 98-100% and 72% sensitivity in smear-negative samples (96). Though the Xpert assay is preferred due to its simplicity, rapidity, higher sensitivity, ease and safety during sample processing and handling, its application in resource-limited countries is hampered by cost (5). Hence, few African countries such as South Africa has implemented Xpert assay as the baseline diagnostic method for pulmonary TB (97).

#### Conventional and real-time PCR

Besides commercial diagnostics, PCR is also used to amplify targeted regions of the *M. tb* genome, which is afterwards sequenced with the ABI PRISM DNA sequencer (Applied Biosystems, Foster City, CA) (Tables 2, S1-S4). Real-time PCR methods however, which is based on hybridization of amplified DNA with fluorescent-labeled probes across DNA regions of interest (78), can detect both TB and its resistance mechanisms (98). The amount of amplified DNA is directly proportional to the fluorescence signal, which can be controlled on-line (20). Different types of probes including TaqMan probe, fluorescence resonance energy transfer probe, molecular beacons, and bioprobes are commonly used (35, 86, 99). However, designing a probe for each single nucleotide polymorphism (SNP) is costly (100); Beacon and TaqMan probes identify commonly encountered SNPs in *katG* and *rpoB*, (20, 55, 101). Fluorescence resonance energy transfer probes can identify multiple SNPs at a cheaper cost, making them ideal for initial screening tests; however, they require two rounds of PCR amplification. (47). Real-time PCR not only allow detection of targeted gene(s) or mutation(s), but also enables quantification of gene expression (55). The other main advantages of real-time PCR methods are their short turnaround time (result can be available 1.5-2.0h after DNA extraction) and low risk of contamination due to its closed single-tube system (20). However, the equipment and reagents are expensive, and the techniques need well-trained personnel; thus, its application in resource-limited countries are limited.

#### WGS

The advantages of WGS in TB diagnosis, typing and resistance detection has informed its adoption as a routine tool in the UK for all suspected TB cases, a feat which is far in the future for Africa (94, 102). Unlike other molecular methods, WGS provides DNA sequence information of the whole genome, enabling the detection of hetero-resistance (both resistant and susceptible subpopulations) in mixed *M. tb* populations, as well as detection of known and previously uncharacterized/unknown gene mutations (103). However, not all gene mutations detected through WGS might confer drug resistance due to hypo- or non-expression (52, 104). Although WGS is expensive and not easily available for routine detection of drug resistance gene mutations, it is the most reliable method to study pleiotropic and polygenic genes that may be involved in resistance to single or multiple drugs (20, 36). Hence, to understand the complete drug resistance determinants in *M. tb* strains, in-depth DNA sequencing (WGS) could be performed (66). However, for mutations concentrated/restricted to short segments of the genome or a gene, such as *rpo*B gene analysis for RIF resistance, PCR-based targeted sequencing is expedient (47). One study reported that WGS improved PZA resistance detection with a sensitivity of 97% and specificity of 94%; WGS was used as gold standard to resolve discordance results from conventional methods (105).

Sanger sequencing is the oldest WGS platform and works by chain termination while Illumina, which is the most common platform in the market currently, uses sequencing by synthesis and is better in its speed and length of readout, producing an overwhelming 1GB of nucleotide per run (106). Pyrosequencing is another sequencing-by-synthesis technique, which is used by the Roche 454 platforms for short read sequencing and has been utilized for *M. tb* drug resistance detection (107). Marttila *et al*. (2009) revealed that the sensitivity of pyrosequencing was 97.4% and 66.7% for accurate detection of RIF and INH resistance, respectively (107). In another study, this technique detected important mutations in the *rpo*B gene in 96.7% of RIF-resistant strains, in *kat*G for 64% of INH-resistant strains, and in *gyr*A for 70% of OFL-resistant isolates (108). Pyrosequencing was preferred over the Sanger method due to its improved turnaround time. However, it is limited by its short-read lengths (up to 20-50 nucleotides) (109). Currently, the Illumina, PacBio and Nanopore sequencers are increasingly being used in well-resourced laboratories to diagnose and type *M. tb* (94).

### Molecular genotyping techniques

#### IS*6110 RFLP*

Molecular genotyping is a vital technique for the understanding of TB epidemiology. These techniques can predict transmission rates and identify dominant strains, strains that stand a chance of spreading and those related to outbreaks (42), severe disease (41), and drug resistance. Evidently, patients with genotypically clustered strains are epidemiologically related, representing recent transmissions (91). The first *M. tb* DNA typing technique to be widely employed was restriction fragment length polymorphism (RFLP). Its standardized protocol was published in 1990 and is currently still used (110). The RFLP technique makes use of IS*6110* insertion sequence (110). There are up to 25 copies of IS*6110* commonly found in almost all strains of *M. tb* complex (111), but not present in any other species. *M. tb* strains are differentiated by looking at the number and position of IS*6110* as it varies between strains. The IS6110 is the current gold standard for *M. tb* complex genotyping, given its superior discriminatory power over other genotyping methods for *M. tb* complex (112).

Isolates from patients infected with epidemiologically unrelated strains of tuberculosis have different RFLP patterns, whereas those from patients with epidemiologically related strains have identical RFLP patterns (112). However, RFLP can only detect strains with fewer than six IS*6110*, necessitating the use of other supplementary genotyping methods (112). Moreover, it is slow, cumbersome, labor-intensive, technically demanding and requires a relatively large amount (2µg) of high-quality DNA. In addition, the interpretation of data can be prone to errors. To get the required amounts of DNA, substantial colonies must be grown from a clinical sample, which can take four to eight weeks of culturing. Furthermore, the technique has poor discriminatory power for isolates with less than six copies of IS*6110*. Further technological refinements resulted in the development of PCR-based methods, which are easier to perform, require relatively smaller amounts of genomic DNA, and can be applied to non-viable bacteria or directly to clinical samples (113).

#### MIRU-VNTR typing

In 2001, a rapid, reproducible typing method based on a variable number of tandem repeats (VNTR), using 12 mycobacterial interspersed repetitive units (MIRU), was developed (114). The MIRU-VNTR method has a resolution power close to that of IS6*110* RFLP when used in typing mycobacterial isolates. MIRU are short (40-100bp) DNA elements found as tandem repeats and dispersed in intergenic regions in the genome of *M. tb* complex (115). It is not yet known whether they are junk DNA, or whether they contribute to biological function within organisms. Strain analysis is done by observing the number of repeats at different loci as they vary per strain. The number of strain loci can influence the interpretation of recent transmission rates, and therefore, an estimation of the rate of clustering; the number of loci that can be used are 12, 15, and 24 (116–118). The type strain is assigned 12 digital numbers corresponding to the number of repeats at each MIRU locus, forming the basis of a coding system that facilitates inter-laboratory comparisons. Moreover, relative to the IS6*110* RFLP typing, MIRU-VNTR profiling is fast, appropriate for specific strains regardless of their IS6*110* copy number and permits rapid comparisons of global strains using a binary data classification system. Recently, a set of 24 MIRU-VNTR loci having higher discriminatory power than the 12-locus system was described, which results in greater performance than the combined resolution of both RFLP and spoligotyping (119, 120). Spoligotyping is a rapid additional typing method used to increase the discriminatory power of MIRU-VNTR typing (121).

#### Spoligotyping

Spoligotyping is a PCR-based typing method that shows the presence or absence of unique spacer sequences, which are found between the direct repeat sequences of the direct repeat (DR) region (113). The DR region contains similar multiple 36bp regions interspersed with non-repetitive spacer sequences of a similar size. As a PCR based method, small amounts of DNA are required. The result of spoligotyping can be expressed in digital format (113, 122).

#### WGS

High throughput genome sequencing technology have an advantage over traditional genotyping methods in distinguishing strains in populations with a high burden of *M. tb* and to assess the rate of transmission in these settings (72, 123). Currently, the molecular typing techniques used for epidemiological and evolutionary applications are IS6*110* RFLP, MIRU-VNTR and spoligotyping. However, recent studies indicate strains with quite similar DNA fingerprinting patterns may harbor considerable genomic diversity (14, 72). Hence, the application of WGS enables detailed understanding of the genetic diversity and phylogeny of drug-resistant *M. tb* complex strains found in certain geographical areas (14, 93, 124). WGS can discriminate relapse TB from re-infection and its resolution has greater advantage over traditional genotyping technologies (93, 100).

Currently, there are different types of NGS platforms with high and moderate throughputs, such as: Illumina (MiSeq, HiSeq, NextSeq and iSeq); Bio-Rad Laboratories (GnuBio); Thermo Fisher (SOLiD, S5, personal genome machine [PGM], Proton); Oxford Nanopore (minION, PromethION, GridION); PacBio (Sequel I, II and RS2); Qiagen (GeneReader); Vela Diagnostics NGS platform (Singapore). Most of these sequencing technologies are used in only high-level (at referral-level) facility, with a few being used in intermediate-level facilities (e.g. the minION, MiSeq and PGM). Although WGS is a highly promising technology in the control of TB disease, it has several drawbacks to its application for clinical use. Few of the challenges are: high amount and quality DNA are required; highly trained staff is needed for sample preparations, processing and sequencing; large amount of raw data generated by NGS and WGS presents challenge to users who are unfamiliar with bioinformatics skills (94).

## 5. Conclusions, limitations and recommendations

The molecular epidemiology, diagnostics, resistance mechanisms and rates in *M. tb* complex strains are herein characterized on a country-by-country basis and in Africa, with INH and RIF resistance being more pronounced in almost all countries. Notably, all the eight *M. tb* lineages were found in Africa, but with different frequencies per country. As well, the clades of *M. tb* strains were more locally based than transnational, suggesting minimal intercountry spread of same clones within Africa. Furthermore, the resistance mutations underlying antibiotic resistance in *M. tb* were myriad, with specific mutation hotspots being commonly found in all lineages and countries on the continent. Obviously limited by skill, cost and accessibility to modern laboratories, the MDR rates in most of the included studies were obtained from a relatively smaller sample size, albeit the values obtained are worrying, particularly in Zimbabwe, Sudan, Swaziland and South Africa. Unfortunately, the XDR rate could not be determined for all the countries except South Africa, further belying the dearth of necessary skill, adequate budget and well-equipped laboratories to diagnose this life-threatening menace.

Advances in genomic engineering and whole-genome sequencing (WGS) techniques are increasing our comprehension of this pathogens’ resistance mechanisms, which is critical for the development of rapid diagnostics and antitubercular drugs (29, 125–127). The limited number of studies using WGS to characterize strains limited the genomic characterization of the resistance mechanisms and epidemiology, particularly as commercial *M. tb* diagnostics and typing tools only target very small regions of the genome. Subsequently, the resistance mutations identified were repetitive as the same primers, probes and diagnostics were used from country to country. Nevertheless, the monoresistance and MDR rates require urgent attention from the respective countries to help contain TB in Africa.

## Data Availability

All data associated with this manuscript is already included in the supplemental files

## Contributors

JOS: Conception, study design, analyses, phylogenomics, images/figure design, writing and formatting manuscript. MAR: literature search, analysis, writing portions of manuscript. NEM and PBF: critical reviewing of manuscript.

## Acknowledgments

None

---

**Figure S1:**
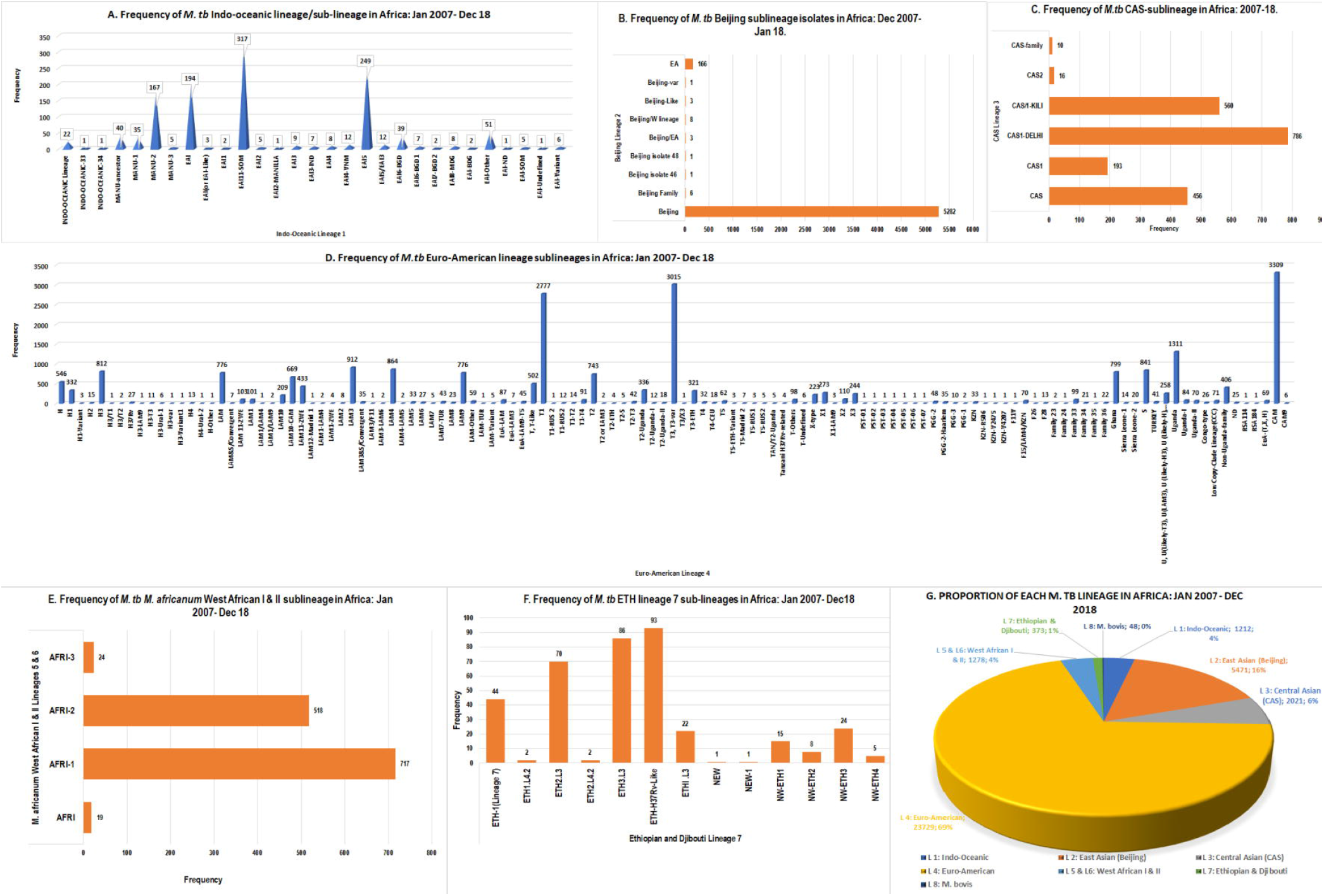
Frequency of *M. tuberculosis* Lineages/sub-lineages in Africa, January 2007-December 2018: Frequency of Indo-oceanic lineage/sub-lineages (**A**); Beijing sublineage (**B**); CAS-sublineage (**C**), Euro-American lineage/sub-lineages (**D**); *M. africanum* West Africa I & II (**E**); *M. tuberculosis* Eth lineage 7 and sub-lineages (**F**); and proportion of each *M. tuberculosis* lineage in Africa (**G**).

**Table S1:** Distribution of *M. tuberculosis* complex strains, specimen source/s, genotyping method/s, molecular anti-TB drug resistance rate and resistance mechanisms in *M. tb* across African countries, January 2007 to December 2018

**Table S2:** Resistance mechanisms, molecular diagnosis method/s used, frequency and proportion of gene mutation per total resistant M. *tuberculosis* complex isolates across African countries, January 2007-December 2018

**Table S3:** Molecular antibiotics resistance rates and resistance mechanisms in *M. tuberculosis* complex across African countries, January 2007-December 2018.

**Table S4:** Frequency of gene mutation(s) and specific amino acid/nucleotide changes conferring antitubercular drug resistances across African countries January 2007-December 2018

**Table S5:** Distribution of specimen source/s, phenotypic DST method/s used, total number of isolates, and phenotypic antibiotics monoresistance rate, MDR and XDR rate of *M. tb* complex across African countries, January 2007-December 2018

**Table S6:** Distribution of genotypes/lineages/sub-lineages, frequency and patterns of antibiotics resistance-conferring mutations across African countries, January 2007-December 2018

**Supplementary dataset 1:** Metadata of *M. tuberculosis* isolates included in phylogenomic analyses.

**Supplementary dataset 2:** Country-by-country frequency of lineage and sub-lineage of *M. tuberculosis* in Africa: January 2007-December 2018

